# Inflammatory arthritis immune related adverse events represent a unique autoimmune disease entity primarily driven by T cells, but likely not autoantibodies

**DOI:** 10.1101/2025.06.06.25328991

**Authors:** Xingxing Zhu, Yue Yu, Yanfeng Li, Panwen Wang, Ying Li, Chantal McCabe, Shiju Chen, Hannah E. Langenfeld, Andrew C. Hanson, Brenna E. Sharp, Amber Woltzen, Cynthia S. Crowson, Svetomir N. Markovic, John M. Davis, Haidong Dong, Uma Thanarajasingam, Hu Zeng

## Abstract

Immune checkpoint inhibitor (ICI)–induced inflammatory arthritis (IA) is an increasingly recognized immune-related adverse event (irAE), yet its underlying immunopathogenesis remains poorly understood. Unlike rheumatoid arthritis (RA), where autoantibodies and B cell dysfunction are central features, the contribution of humoral immunity to IA irAE is unclear. Here, we performed in-depth immunophenotyping of peripheral blood from patients with IA irAE, and compared them with seronegative RA patients, ICI-treated patients without irAE, and healthy controls. IA irAE was marked by a distinct T cell–dominated profile, with CD4⁺ T cells exhibiting reduced CXCR3 and CCR6 expression, and both CD4⁺ and CD8⁺ T cells showing increased cytotoxic molecule expression and metabolic activation. In contrast to seronegative RA, IA irAE patients displayed no significant elevation in autoantibody levels or atypical CD11c⁺CD21⁻ B cells. IA irAE was further characterized by a proinflammatory cytokine milieu, with elevated levels of IL-6, IL-12, and type I IFN, which correlated with the observed T cell activation phenotypes. Altogether, our findings define IA irAE as an immunologically distinct entity from RA, representing a naturally occurring model of antibody-independent systemic autoimmunity in humans. These results suggest that pathogenic T cell responses, potentiated by specific inflammatory cytokines, may be sufficient to drive arthritis in the absence of humoral autoimmunity, offering new insights into immune tolerance breakdown and therapeutic targeting in irAEs.

**One Sentence Summary:** inflammatory arthritis irAE is a T cell driven, autoantibody independent autoimmunity

## INTRODUCTION

While the revolutionary immune checkpoint inhibition (ICI) therapy invigorates anti-tumor immunity, it also leads to various immune related adverse events (irAEs). Rheumatic irAEs account for about 5% of all irAEs. Most of the rheumatic irAEs are considered inflammatory arthritis (IA) (*1, 2*). From an immunological point of view, and based on genetic mouse models, the development of irAEs could be considered an expected outcome because ICI therapy blocks key inhibitory receptors, CTLA-4 or PD-1. CTLA-4 deficiency leads to severe systemic autoimmunity and early death (*3*), whereas PD-1 deficiency results in autoimmune diseases that vary in onset, severity and affected tissues depending on genetic background (*4–6*). Antibodies targeting PD-1 or its ligands, PD-L1, are the most used ICI agents in clinical practice. So far, the immunological underpinnings of the irAE development remain elusive. Furthermore, while it is widely assumed that ICI therapy primarily targets CD8^+^ T cells, PD-1 is expressed and functional on CD4^+^ T cells and non-T cells, including B cells (*7*). Yet, its impact on patient humoral immunity remains contentious.

The impact of PD-1 inhibition on humoral immunity has been mostly explored in the context of vaccination. While some studies showed that anti-PD-1/PD-L1 agents do not significantly affect the titer of anti-SARS-CoV2 spike protein antibodies following COVID-19 vaccination (compared to non-anti-PD-1/PD-L1 treatment) (*8–11*), one study found that anti-PD-1/PD-L1 treatment is associated with significantly increased post COVID-19 vaccination SARS-CoV2 breakthrough infection (*12*). Intriguingly, an elegant influenza vaccination study found that anti-PD-1 treatment reduces the quality of the influenza-specific antibodies (*13*). Importantly, a genetic study on individuals with complete loss of PD-1 demonstrated that B cell intrinsic PD-1 is critical for memory B cell formation and the production of antibodies against common microbial pathogens (*14*). Finally, one study found that anti-PD-1/PD-L1 treatment is associated with reduced autoantibody profile compared to anti-CTLA-4 treatment (*15*). Thus, existing data suggest that anti-PD-1/PD-L1 treatment might attenuate vaccine elicited immune protection, and B cell intrinsic PD-1 may enhance humoral immunity.

Previous studies using relatively small patient cohorts have revealed that CD8^+^ T cells from IA irAE patients have elevated glucose metabolism at baseline (*16*), and there is a clonal expansion of CD38^hi^ cytotoxic, CX3CR1^+^ effector CD8^+^ T cells associated with elevated expression of type I interferon (IFN) inducible genes, and increased Th1/2-like T cell transcription signatures in these patients (*17–20*). Yet, several important questions remain, including any biomarkers distinguishing ICI treated patient without irAE from those with IA irAE, the involvement of humoral immunity, and the relationship between IA irAE and rheumatoid arthritis (RA) with shared clinical and serological phenotypes.

It has been well recognized that systemic breakdown of self-tolerance during the pathogenesis of RA usually involves both autoreactive T cells and autoreactive B cells, which generate autoantibodies, often with T cell help (*21, 22*). Autoantibodies may lead to arthritis by forming immune complexes (ICs) to promote complement deposition in joint tissues. Yet, there is growing evidence that inflammatory arthritis could be antibody-independent because complement fixation or ICs can be absent in some patients with inflammatory arthritis (*23*).

Animal models further demonstrate the possibility of T cell driven and B cell independent arthritis, including SKG mice (*24*) and IL-1ra deficient mice (*25*). Nonetheless, the existence of antibody-independent arthritis in humans remains a contentious issue. The immunological basis for possible T cell mediated antibody-independent arthritis in humans is poorly defined.

Here, we perform comprehensive immunological analyses, integrating single cell RNA sequencing, flow cytometry, proteomics, multiplex assays and cell culture on a large cohort of inflammatory arthritis irAE patients, with sex, age and serologically matched RA patients, ICI treated cancer patients without irAE and healthy controls. Our data demonstrated that inflammatory arthritis irAE is immunologically distinct from RA, associated with increased cytotoxic T cell signatures, metabolic activities, reduced expression of CXCR3 and CCR6 on CD4^+^ T cells, and highly elevated levels of multiple cytokines and chemokines, but notassociated with increased autoantibody production or pathological atypical B cells. These phenotypes are partly driven by increased IL-6, IL-12 and type I IFN. Thus, our results indicate that IA-irAE is potentially a T cell driven and antibody independent disease entity immunologically distinct from conventional RA.

## RESULTS

### Patient characteristics

From 2017 to 2024, we enrolled and performed analysis on 171 patients, who were divided into 4 groups, rheumatic irAE (irAE, n = 34), ICI control (ICI, n = 26), RA control (RAC, n = 47), healthy control (HC, n = 64) (Summarized in **Supplementary Table 1**). The average age of the patients in our cohort was 62. In the irAE and ICI control groups, the majority were male (58.8% and 57.7%, respectively), while in the RA control group, males were a minority (38.3%). Both irAE and ICI control patients had a mixture of different cancer diagnoses, and most (97.1% for irAE, 88.5% for ICI) were at tumor stage IV. In terms of immune checkpoint inhibition medicine, the majority of patients with either irAE (97.1%) or ICI control (88.5%) were treated with anti-PD-1/PD-L1 therapy. Following the immunotherapy treatment, all the patients in the irAE group developed inflammatory arthritis (100%, 34/34), while other irAEs included systemic lupus erythematosus (2.9%, 1/34) and antisynthetase syndrome (2.9%, 1/34). The majority of the irAE patients received steroid treatment (76.5%, 26/34). After ICI treatment, more cancer patients in irAE group had a complete response than those in ICI control group (20/34, 58.8% vs 9/26, 34.6%, p = 0.0742), indicating a possible association with irAE development and better tumor response to treatment. Lastly, serological tests showed that the majority of irAE patients are negative for rheumatoid factor (RF) (3/31, 9.7%) and anti-cyclic citrullinated peptide antibody (anti-CCP) (2/30, 6.7%). All RF^+^ and anti-CCP^+^ irAE patients had relatively low titers of RF or anti-CCP (**Supplementary Table 1**). Therefore, most of the irAE patients were considered seronegative, consistent with previous studies (*19, 26–28*).

### CD8^+^ T cells from irAE patients have increased cytotoxic T cell features

To define molecular and cellular changes associated with the development of clinical arthritis after immunotherapy, we performed single-cell RNA sequencing (scRNAseq) analysis on the peripheral blood mononuclear cells (PBMCs) from irAE patients, RA control patients, ICI control patients, and healthy controls. After stringent quality control and filtering steps, approximately 220,000 cells were analyzed with respect to their transcriptomes. We performed supervised cell subset annotation according to the literature (*29, 30*). Uniform Manifold Approximation and Projection (UMAP) illustrated major immune cell subsets, including B cells, CD4^+^ T cells, CD8^+^ T cells, dendritic cells (DC), monocytes and natural killer cells (NK) (fig. S1A). scRNAseq showed that the proportion of CD8^+^ effector memory cell re-expressing CD45RA (TEMRA) was increased in irAE and RA patients compared to HC and ICI control, meanwhile both irAE and ICI control exhibited higher CD8^+^ TEM compared to HC and RA control (Fig. 1A). Flow cytometry analysis showed that both irAE and ICI control patients had higher CD45RA^+^CCR7^-^ population relative to HC and RA control, although the increase was only significant in irAE patients (Fig. 1B). The discrepancy in terms of TEMRA in ICI controls between scRNAseq and flow cytometry may be because sequencing analysis used a set of 21 genes to annotate TEMRA whereas flow cytometry relied on 2 markers and thus might miss the heterogeneity of CD45RA^+^CCR7^-^ population.

**Fig. 1.**
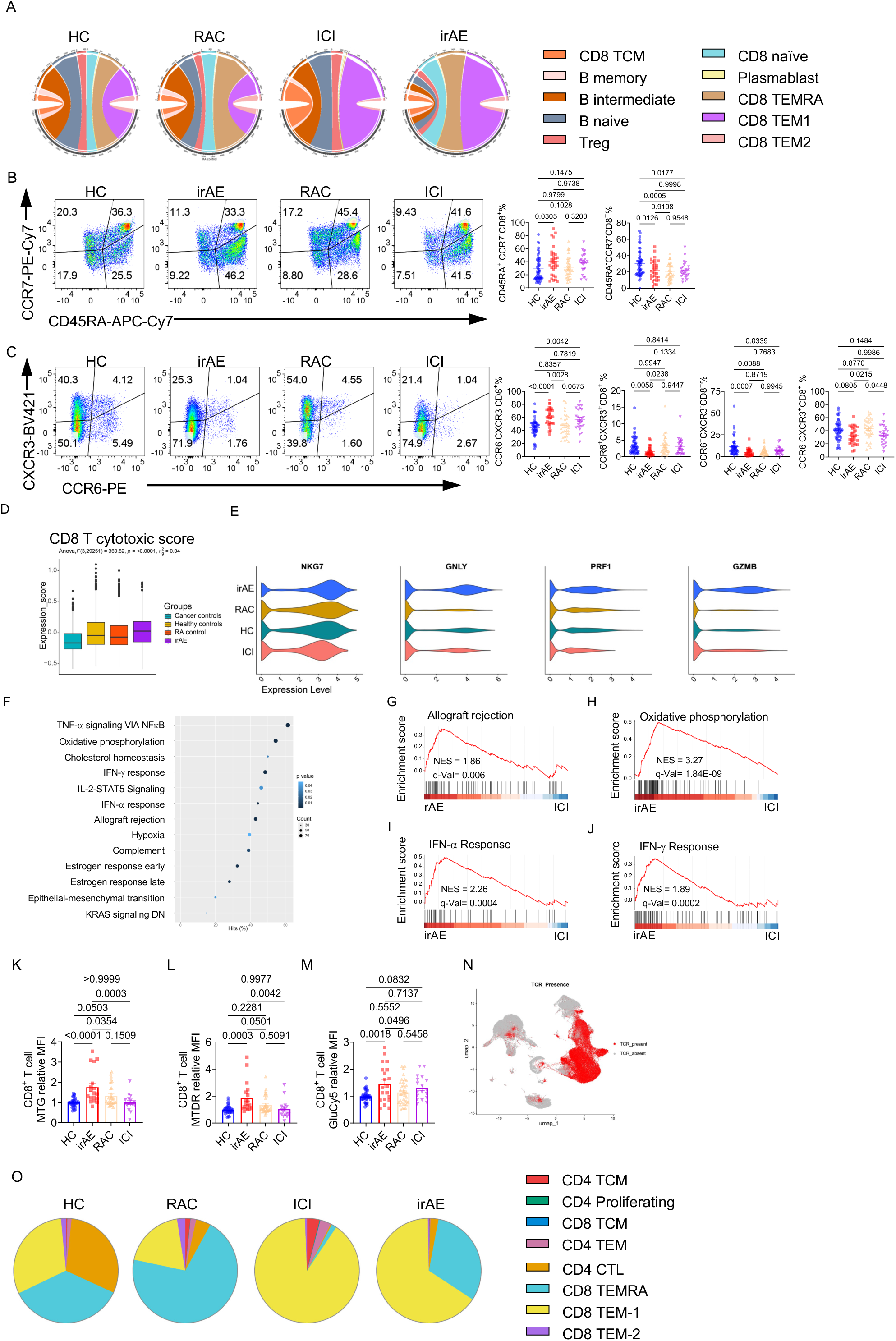
CD8^+^ T cells from the irAE patients are more cytotoxic and metabolic active. (A) Circos plots showing the percentage of different subsets of CD8^+^ T cells and B cells from HC (Healthy control, n = 6), ICI (ICI control, n = 2), RAC (RA control, n = 5), or irAE (n = 5). (B) Expression of CD45RA and CCR7 on CD8^+^ T cells. Right, summaries of the percentage of CD45RA^+^CCR7^-^CD8^+^ T cells and CD45RA^-^CCR7^-^CD8^+^ T cells from HC (n = 53), irAE (n = 29), RAC (n = 41), ICI (n = 26). (C) Expression of CXCR3 and CCR6 on CD8^+^ T cells. Right, summaries of CXCR3^-^CCR6^-^CD8^+^, CXCR3^+^CCR6^+^CD8^+^, and CCR6^+^CXCR3^-^CD8^+^ T cells. HC (n = 45), irAE (n = 27), RAC (n = 31), ICI (n = 26). (D) The cytotoxic score was evaluated on CD8^+^ T cells using the gene list identified previously (*86*). (E) Specific cytotoxic or effector function-related genes were evaluated on the CD8^+^ T cells. (F-J) GSEA was performed on the CD8^+^ T cells between irAE and ICI. (F) Pathways that were significantly enriched in the comparison. GESA plots of the allograft rejection (G), oxidative phosphorylation (H), IFN-α response (I), and IFN-γ response (J) between irAE and ICI. (K-M) PBMCs from different groups were stimulated with 10 μg/mL plate-coated anti-CD3 and anti-CD28 for 5 days. Mean fluorescence intensities (MFI) of mitochondria green (MTG) (K), mitochondria deep red (MTDR) (L) (HC (n = 31), irAE (n = 18), RAC (n = 32), ICI (n = 16)) or Cy5-linked-1-amino-glucose (GluCy5) (M, HC (n = 35), irAE (n = 20), RAC (n = 36), ICI (n = 18)) in CD8^+^ T cells were presented. Expression was normalized to the HC in each experiment. (N) UMAP shows the presence or absence of TCR in the major immune cells across all the samples. (O) Pie charts showing the distribution of the top 100 TCR clones across different T cell subsets within each group. Data in graphs represent mean ± SEM, Significance was tested by one-way ANOVA.

Central memory CD8^+^ T cells (CD45RA^-^CCR7^+^) and naïve CD8^+^ T cells (CD45RA^+^CCR7^+^) were unaltered among groups (fig. S1B). Further analysis revealed that CD45RA^+^CCR7^-^CD8^+^ TEMRA cells had the highest expression of NKG7, a gene critical for T cell cytotoxicity (*31*) (fig. S1C). We also observed distinct chemokine receptor expression in irAE and ICI patients, both had increased proportion of CCR6^-^CXCR3^-^CD8^+^ cells and reduced proportion of CCR6^+^CXCR3^+^CD8^+^ T cells compared to HC or RA controls (Fig. 1C), implying that PD-1/PD-L1 blockade reduces CXCR3 and CCR6 expression on CD8^+^ T cells. Flow cytometry analysis indicated that CCR6^-^CXCR3^-^CD8^+^ T cells from healthy donors contained a higher proportion of CD45RA^+^CCR7^-^CD8^+^ T cells (fig. S1D), and with relatively higher NKG7 expression (fig. S1E). Moreover, RNAseq analysis revealed that CCR6^-^CXCR3^-^CD8^+^ T cells from healthy donors had higher expression of TEMRA signature genes (*29*) (fig. S1F), and were enriched with natural killer cell cytotoxicity (fig. S1, G and H), relative to CCR6^+^CXCR3^-^ and CCR6^-^CXCR3^+^ CD8^+^ T cells. Further, effector T cell associated markers, including the proportion of CD28^-^CD8^+^ T cells (fig. S1I), CD38^+^CD127^-^CD8^+^ T cells (fig. S1J), and expression of a terminal effector T cell marker CX3CR1 (fig. S1K) were all increased in both irAE patients and ICI control relative to HC and RA controls, although the increases were more consistent in irAE than in ICI controls.

These markers were interlinked because CD38^+^CD127^-^ cells were significantly enriched in CX3CR1^+^CD8^+^ T cells relative to CX3CR1^-^CD8^+^ T cells (fig. S1L), and CD28^-^CD8^+^ T cells had higher expression of CX3CR1 and a higher proportion of CD38^+^CD127^-^ cells than CD28^+^CD8^+^ T cells (fig. S1, M and N). Furthermore, all these biomarkers were also higher in CXCR3^-^CCR6^-^ CD8^+^ T cells (fig. S1, O-Q), indicating an inverse correlation between chemokine receptors, CXCR3, CCR6, and CX3CR1 (*32, 33*). Thus, our data indicate that while CD8^+^ T cells from both irAE and ICI control patients exhibit increased expression of some effector T cell markers, the changes were more consistent in irAE patients.

Compared to ICI controls, IrAE CD8 T cells had higher expression of TEMRA signatures that include many cytotoxic genes, suggesting an increased expression of the cytotoxic T cell program. Indeed, bioinformatic analysis showed irAE CD8^+^ T cells had the highest cytotoxicity score (Fig. 1D) and the highest expression of cytotoxic molecules, such as NKG7, GNLY, PRF1 and GZMB (Fig. 1E). Gene set enrichment analysis (GSEA) showed that irAE CD8^+^ T cells were enriched with genes involved in TNF signaling, IFN-α response, IFN-γ response, allograft rejection, and oxidative phosphorylation compared to ICI controls (Fig. 1F-1J). Consistent with the enriched oxidative phosphorylation related genes, irAE CD8^+^ T cells had highest mitochondrial mass and mitochondrial membrane potential indicated by mitotracker green (MTG) and mitotracker deep red (MTDR) stainings, respectively (Fig. 1K and 1L), suggesting possibly higher mitochondrial activity in irAE CD8^+^ T cells. Additionally, irAE CD8^+^ T cells had highly increased glucose uptake as measured by Cy5-linked-1-amino-glucose (GluCy5) staining (*34*), suggesting an increased glucose metabolism (Fig. 1M). Finally, TCR repertoire analysis showed that there was a clonal expansion of CD8 TEM or TEMRA cells in irAE patients (Fig. 1N and 1O). Altogether, our data indicate that CD8^+^ T cells from irAE patients are metabolically active and functionally cytotoxic, which distinguishes them from all control patients.

### CD4^+^ T cells in the irAE patients are metabolically active and have increased CXCR3^-^ CCR6^-^ frequency and reduced CXCR3^+^CCR6^+^ frequency

Consistent with previous literature (*35–37*), the CD4/CD8 ratio increased in the RA control patients, but it remained unaltered in irAE patients and ICI controls (Fig. 2A). Within CD4^+^ T cells, a significant decrease of the CD127^lo^CD25^+^CD4^+^ regulatory T cells (Treg) was noted in RA patients, but not in irAE or ICI controls (Fig. 2B), suggesting that ICI therapy did not significantly affect circulating Treg frequency. Human circulating CXCR5^+^CD4^+^ T cells have B cell helper functions analogous to follicular helper T cells (Tfh) (*38–40*). The proportion of CXCR5^+^CD45RA^-^CD4^+^ T cells significantly decreased in the irAE and ICI control patients compared to HC (Fig. 2C). However, this reduction was only significant in irAE patients, but not ICI, compared to RA patients (Fig. 2C). Within CXCR5^+^CD45RA^-^CD4^+^ T cells, the proportion of CXCR3^+^CCR6^-^, CXCR3^-^CCR6^+^, and CXCR3^+^CCR6^+^ populations remained unaltered among patient groups (fig. S2A). To further investigate other T cell lineages, we use surface molecules CXCR3 and CCR6 to interrogate Th1-like (CXCR3^+^CCR6^-^), Th17-like (CXCR3^-^CCR6^+^), Th1/Th17-like (CXCR3^+^CCR6^+^), and CXCR3^-^CCR6^-^ cells (*41–43*). The proportion of Th1/Th17-like cells was significantly reduced in irAE patients compared to HC, RA, and ICI controls, while CXCR3^-^CCR6^-^ cells increased in irAE patients relative to HC, RA and ICI controls (Fig. 2D).

**Fig. 2.**
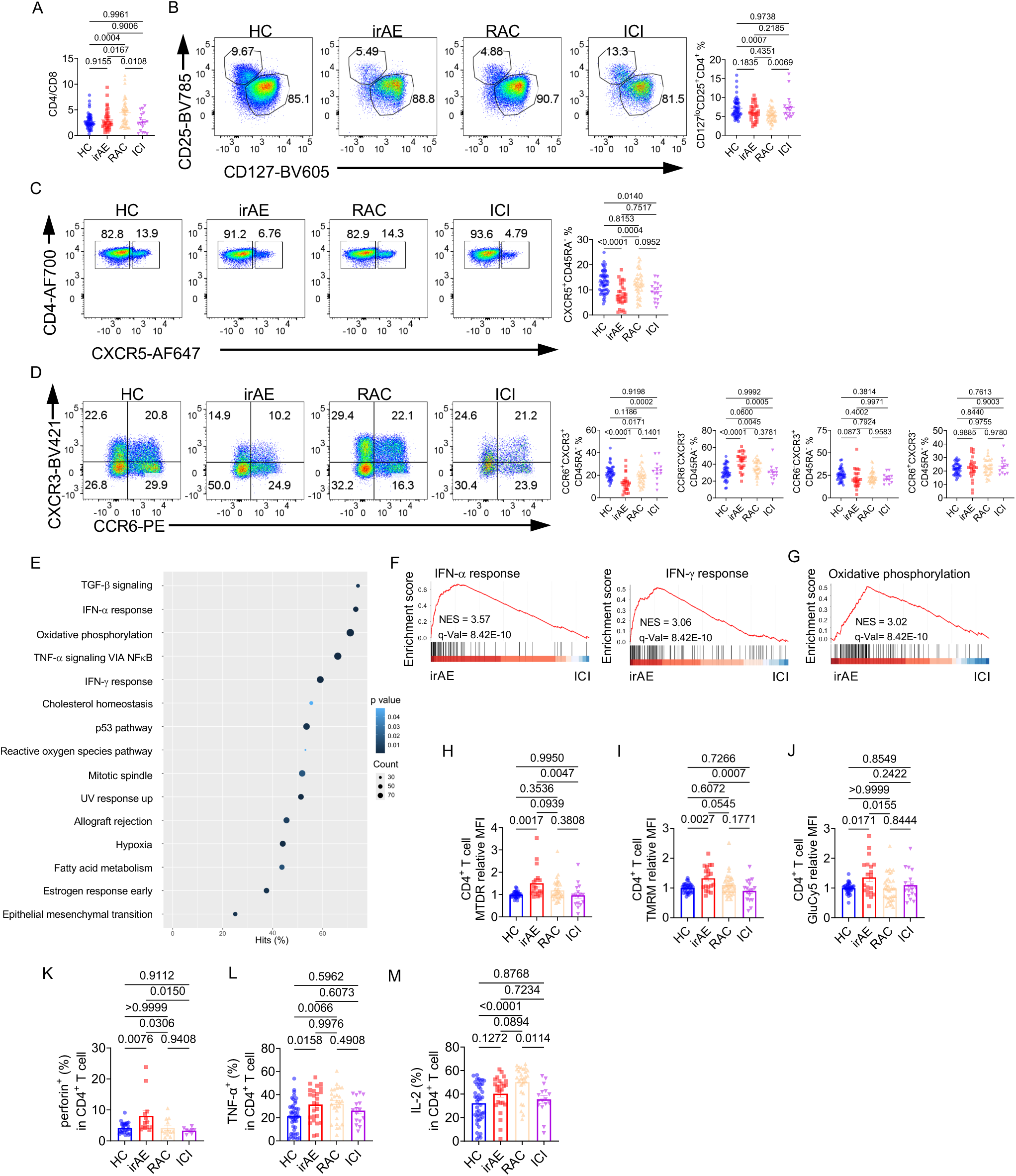
CD4^+^ T cells are metabolic active and have reduced CXCR3 and CCR6 expression in irAE patients (A) The ratio of CD4 to CD8 in PBMCs was calculated. HC (n = 53), irAE (n = 23), RAC (n = 42), ICI (n = 17). (B) Expression of CD25 and CD127 on CD4^+^ T cells. Right, percentage of CD25^+^CD127^lo^ regulatory T cells in CD4^+^ T cells, HC (n = 53), irAE (n = 27), RAC (n = 42), ICI (n = 17). (C) Expression of CXCR5 and CD4 on the CD45RA^-^CD4^+^ T cells. Right, percentage of CXCR5^+^CD45RA^-^CD4^+^ T cells, HC (n = 53), irAE (n = 27), RAC (n = 42), ICI (n = 17). (D) Expression of CCR6 and CXCR3 on CD45RA^-^CD4^+^ T cells. Right, percentages of CXCR3^+^CCR6^-^CD45RA^-^, CXCR3^+^CCR6^+^CD45RA^-^, CXCR3^-^CCR6^-^CD45RA^-^ CD4^+^ T cells, HC (n = 41), irAE (n = 26), RAC (n = 36), ICI (n = 12). (E-G) GSEA was performed on the CD4^+^ T cells between irAE and ICI. (E) Significantly enriched pathways in CD4^+^ T cells from irAE and ICI. GSEA plots of IFN-α and IFN-γ response (F), and oxidative phosphorylation (G). (H-M) PBMCs from different groups were stimulated with 10 μg/mL plate-coated anti-CD3 and anti-CD28 for 5 days, Mean fluorescence intensities (MFI) of MTDR (H, HC, n = 31; irAE, n = 18; RAC, n = 32; ICI, n = 16), TMRM (I, HC, n = 34; irAE, n = 19; RAC, n = 35; ICI, n = 17), or GluCy5 (J, HC, n = 35; irAE, n = 20; RAC, n = 36; ICI, n = 18) in CD4^+^ T cells were presented. Expression was normalized to the HC in each experiment. (K-M) Fresh PBMCs from different groups were stimulated with PMA/Ionomycin/Monensin for 5 h, and the frequencies of perforin^+^ (K, HC, n = 27; irAE, n = 13; RAC, n = 12; ICI, n = 8), TNF-α^+^ (L) and IL-2^+^ (M) ( HC, n = 44; irAE, n = 25; RAC, n = 27; ICI, n = 16) CD4^+^ T cells were examined. Data in graphs represent mean ± SEM, Significance was tested by one-way ANOVA.

Thus, irAE development is uniquely associated with increased CXCR3^-^CCR6^-^ CD4^+^ T cell frequency and reduced CXCR3^+^CCR6^+^ Th1/Th17-like cell frequency. Anti-PD-1/PD-L1 therapy may reduce circulating Tfh-like cell frequency regardless of irAE development.

GSEA identified significantly altered pathways between irAE and ICI patients, including IFN-α response, IFN-γ response, oxidative phosphorylation, hypoxia, TGF-β signaling, and fatty acid metabolism (Fig. 2E). IFN-α response, IFN-γ response, and oxidative phosphorylation pathways were enriched in CD4^+^ T cells from irAE patients (Fig. 2F and 2G). Moreover, IFN-α response and IFN-γ response were also significantly enriched in irAE CD4^+^ T cells compared to RA controls (fig. S2B) or HC (fig. S2C). CD4^+^ T cells from irAE patients had higher mitochondrial membrane potential measured by MTDR (Fig. 2H) and tetramethylrhodamine methyl ester (TMRM) (Fig. 2I), increased mitochondrial mass measured by MTG (fig. S2D), increased glucose uptake measured by GluCy5 staining (Fig. 2J), and increased reactive oxygen species measured by CellRox (fig. S2E), indicating an overall heightened cellular metabolism. Elevated glucose-dependent metabolism in irAE CD4^+^ T cells was confirmed by SCENITH assay, which uses the energy-intensive protein-synthesis process (measured by puromycin incorporation) as a readout of metabolic activities (fig. S2F). Consistent with higher metabolic signatures in irAE CD4^+^ T cells, RNA sequencing results showed that healthy donor CXCR3^-^CCR6^-^ CD4^+^ T cell subset was enriched in genes associated with energy intensive ribosomal biogenesis and translation program (fig. S2, G-I). Glucose metabolism is closely linked to the effector/cytotoxic CD4^+^ T cell function (*44, 45*). CD4^+^ T cells from irAE patients had the highest perforin expression among the 4 groups (Fig. 2K), consistent with the observations in other autoimmune diseases (*46, 47*) and suggesting increased cytotoxic activity in irAE CD4^+^ T cells. TNF-α production was increased in both irAE and RA CD4^+^ T cells compared to HC (Fig. 2L). Notably, IL-2 was highly increased on CD4^+^ T cells from RA patients, but not irAE and ICI patients (Fig. 2M), suggesting a disease specific cytokine expression pattern in CD4^+^ T cells. Overall, CD4^+^ T cells from irAE patients were metabolically active, exhibited enhanced effector and cytotoxic functions, and were distinguished by reduced expression of CXCR3 and CCR6.

### The humoral immune compartment remains largely unaltered in irAE patients

It has been established that both T cells and B cells play critical roles in the pathogenesis of RA (*21*). To gain insight into the B cell compartment in irAE patients, we analyzed the PBMCs from the patients using flow cytometry (gating strategy was shown in fig. S3A). Total B cell (CD19^+^CD14^-^CD3^-^) frequency trended lower, although not statistically significant, in irAE patients relative to healthy donors (fig. S3B). We did not observe significant differences in naïve B cells (IgD^+^CD24^-^CD27^-^CD38^-^CD19^+^, Fig.3A; IgD^+^CD27^-^ CD19^+^, fig. S3C), CD27^+^CD38^+^ antibody-secreting cells (ASC, Fig. 3B), including CD138^+^ ASC or CD138^-^ ASC (Fig. 3C), IgD^-^ CD27^+^ memory B cells (fig. S3D), and CD27^+^CD38^-^ memory B cells between irAE and other groups (Fig. 3B). Double-negative (DN) B cells have been implicated in the development of various autoimmune diseases including rheumatoid arthritis (RA) (*48, 49*). Two strategies for defining DN B cells were used (*50–53*) (fig. S3A). The frequency of DN B cells was not altered in irAE patients using either gating strategy (Fig. 1A and fig. S3E). However, the atypical DN B cells (CD19^+^CD11c^+^CD21^-^) were significantly elevated only in RA patients (Fig. 3D). A significant increase of CD21^-^CD19^+^, and CD11c^+^CD21^-^IgD^-^CD27^-^ B cells (fig. S3F) was also observed in RA patients compared to HC. Other B cell subsets including transitional B cells (*54*) (CD24^+^CD38^+^CD27^-^CD19^+^, fig. S3G), IgD^+^CD27^+^ or IgM^+^IgD^+^CD27^+^CD38^-^ unswitched memory B cells (fig. S3H) all remained largely intact among patients. Moreover, GSEA showed that several pathways were enriched in irAE B cells relative to ICI control (Fig. 3E), including IFN-α response and oxidative phosphorylation (Fig. 3F and 3G). These data indicate that inflammatory arthritis irAE may not be associated with any significant alterations in the peripheral B cell subsets, while RA is associated with significantly increased CD11c^+^CD21^-^ atypical B cells.

**Fig. 3.**
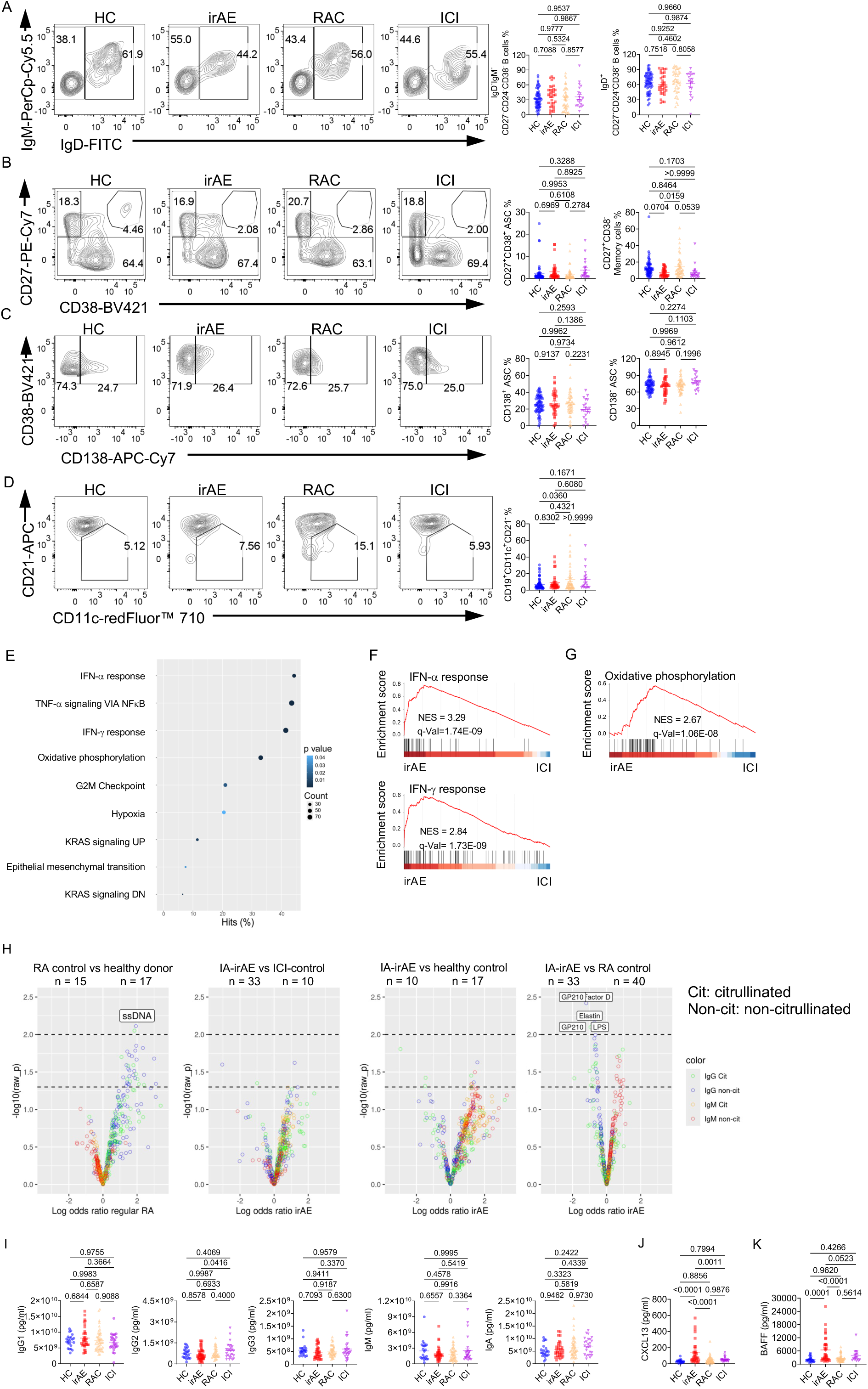
Humoral immunity remains largely intact in irAE patients, while seronegative RA patients have increased atypical B cell frequency and autoantibodies. (A) Expression of IgM and IgD on CD27^-^CD24^-^CD38^-^ B cells. Right, percentage of IgD^+^CD27^-^ CD24^-^CD38^-^ naïve B cells and IgM^-^IgD^-^CD27^-^CD24^-^CD38^-^ double-negative B cells. (B) Expression of CD27 and CD38 on CD19^+^ B cells. Right, percentages of CD27^hi^CD38^hi^ antibody-secreting cells (ASCs) and CD27^+^CD38^-^ memory B cells. (C) Expression of CD138 on CD27^hi^CD38^hi^ ASCs. Right, percentage of CD138^+^ ASCs and CD138^-^ ASCs. (D) Expression of CD11c and CD21 on CD19^+^ B cells. Right, percentage of CD11c^+^CD21^-^CD19^+^ B cells. A-D, HC (n = 63), irAE (n = 33), RAC (n = 46) ICI, (n = 20). (E-G) GSEA was performed on the B cells between irAE and ICI. (E) Significantly enriched pathways in B cells from irAE and ICI. GSEA plots of IFN-α and IFN-γ response (F), and oxidative phosphorylation (G). (H) The volcano plot of the citrullinated or non-citrullinated relative IgG or IgM-isotype autoantigen levels comparing RA vs HC, irAE vs ICI, irAE vs HC, or irAE vs RA. The autoantigens were labeled when p < 0.01. (I) Immunoglobulin isotype levels in the plasma were measured by multiplex assay, HC (n = 22), irAE (n = 34), RAC (n = 46), ICI (n = 26). (J) CXCL13 levels in the plasma were measured by ELISA, HC (n = 20), irAE (n = 34) RAC (n = 47), ICI (n = 23). (K) BAFF levels in the plasma were measured by multiplex assay, HC (n = 21), irAE (n = 34), RAC (n = 47), ICI (n = 18). Data in graphs represent mean ± SEM, Significance was tested by one-way ANOVA (A-D, I-K), logistic regression (H).

Since CD11c^+^CD21^-^ atypical B cells are known to produce autoantibodies and contribute to the development of systemic autoimmunity (*52, 55, 56*), we performed a 128-plex autoantigen array using patient plasma samples on 33 irAE patients, 40 RA-, 10 ICI-, and 17 healthy-controls.

Compared to the healthy control, RA patients (n = 15 females, age < 64 years) had significantly increased levels of multiple IgG autoantibodies, both citrullinated and non-citrullinated, some of which targeted ssDNA, complement C3, U1-snRNP, and TNF-α (Fig. 3H, and fig. S3I). These observations were consistent with previous studies that seronegative RA patients exhibit increased autoantibodies, including ACPA fine specificities, even though they were negative for anti-CCP or RF in standard clinical tests (Supplementary Table 1), highlighting the presence and possible contribution of autoantibodies in seronegative RA patients (*57, 58*). In contrast, we observed few substantial increases of either citrullinated or non-citrullinated IgG isotypes in irAE patients compared to all other control cohorts, although irAE patients had some slightly (but mostly nonsignificant) increased IgM autoantibodies compared to ICI and HC controls (n = 10 female irAE patients age < 64 years for comparison with HC) (Fig. 3H, and fig. S3I).

Interestingly, irAE patients had significantly reduced IgG autoantibodies, such as anti-factor D, GP210, LPS and elastin, compared to RA controls (Fig. 3H, and fig. S3I). Furthermore, because overall immunoglobulin isotype concentrations remain comparable between all 4 cohorts of patients (Fig. 3I), the increased autoantibodies could be the consequence of specific autoreactive B cells in the RA control patients. Hence, our results suggest that IA-irAE might be a unique subtype of arthritis distinct from seronegative RA, characterized by its lack of strong B cell activation and IgG isotype autoantibody signatures. In contrast, seronegative RA is associated with increased atypical B cells and production of IgG autoantibodies.

We conducted a further analysis of several cytokines associated with B cells. B cell chemoattractant CXCL13, and B cell survival factor, BAFF, were uniquely elevated in irAE patients (Fig. 3J and 3K), whereas sCD40L and April were comparable across 4 cohorts of patients (fig. S3J). These data suggest that plasma CXCL13 and BAFF levels might not accurately reflect the activity of autoreactive B cells, but could be associated with irAE development.

### PD-1 blockade does not substantially affect B cell activation and immunoglobulin production *ex vivo*

To gain insight into the effect of PD-1 blockade specifically on B cells, we activated human naïve B cells with different TLR ligands or anti-CD40 in the presence of isotype control or pembrolizumab (Keytruda). Pembrolizumab showed no significant impact on human naïve B cells activation as indicated by the percentage of CD27^+^CD38^+^, CD27^+^CD38^-^ (Fig. 4A) and CD138^+^CD27^+^CD38^+^ B cells (Fig. 4B), as well as the B cell activation marker CD86 (fig. S4A).

**Fig. 4.**
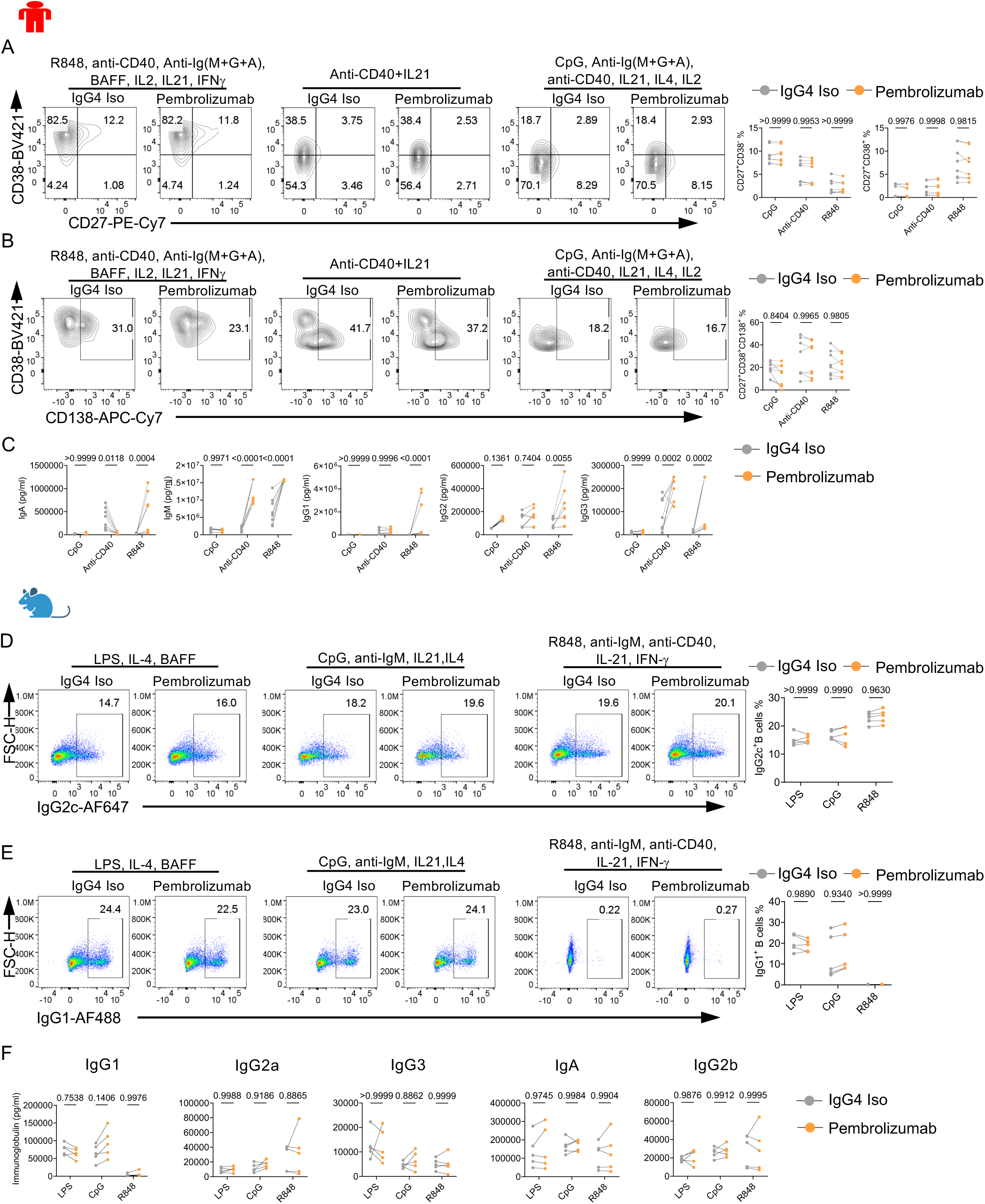
Pembrolizumab does not substantially affect B cell activation and antibody production *ex vivo* (A-C) Human naïve B cells were isolated from PBMCs and cultured under indicated conditions for 7 days; Pembrolizumab or isotype control IgG4 was added on Day 2, n = 8. (A) Expression of CD38 and CD27 on B cells. Right, percentages of CD27^+^CD38^-^, CD27^+^CD38^+^ B cells. (B) Expression of CD138 on CD27^+^CD38^+^ B cells. Right, percentage of CD138^+^CD27^+^CD38^+^ B cells. (C) Different immunoglobulin isotypes were measured in the culture supernatant of (A-B) by multiplex assay. (D-F) B cells from HuPD-1 mice were isolated and cultured with LPS, IL4, BAFF, or ODN2006, anti-IgM, IL-21, IL-4 or R848, anti-IgM, anti-CD40, IL-21, IFN-γ for 3 days, pembrolizumab or isotype control was added on Day 1. (D) Expression of IgG2c on activated B cells. Right, percentage of IgG2c^+^ B cells, n = 5. (E) Expression of IgG1 on activated B cells. Right, percentage of IgG1^+^ B cells, n = 5. (F) Different immunoglobulin isotypes in the supernatant of were measured by multiplex assay, n = 5. Data in graphs represent mean ± SEM, Significance was tested by Two-way ANOVA.

The impacts of pembrolizumab on immunoglobulin production appeared to depend on the type of stimuli. For example, IgA production was reduced by pembrolizumab under anti-CD40 but increased under TLR7/8 ligand R848. However, none of the immunoglobulins were affected by PD-1 blockade under TLR9 ligand CpG (Fig. 4C). Yet, under an age-associated B cells (ABC) skewing condition (R848, anti-Ig(M+A+G), anti-CD40, rhIL-2, rhIL-21, BAFF IFN-γ) (*59*), pembrolizumab increased the production of all immunoglobulin isotypes. These data were largely consistent with a recent publication using PD-1 deficient patient B cells (*14*) and indicated a context dependent effect of PD-1 blockade on antibody production.

Furthermore, we utilized a humanized PD-1/PD-L1 mouse model, in which the endogenous mouse exons coding for the extracellular domains of PD-1 (exons 2 and 3) or PD-L1 (exons 3 and 4) were replaced with the coding sequence for the corresponding human exons. It enabled the testing of FDA-approved PD-1 blocking antibodies on mice (*60*). As expected, pembrolizumab treatment efficiently blocked the detection of surface PD-1 (fig. S4B). Yet, pembrolizumab did not affect the expression of IgG1^+^, IgG2c^+^ (Fig. 4D and 4E), CD138^+^ (fig. S4C), upregulation of activation markers CD69 and CD86 (fig. S4D), metabolic marker CD71 and CD98 (fig. S4E), cell proliferation (fig. S4F), as well as the production of immunoglobin levels in the cultural supernatant (Fig. 4F). Together, these data indicate that PD-1 blockade may not substantially affect B cell activation. However, pembrolizumab may modulate B cell immunoglobulin production in a context-dependent manner.

### Inflammatory cytokines enriched in the plasma of irAE patients may negatively regulate B cell antibody production

To assess soluble factors associated with autoimmunity, we measured plasma concentrations of common cytokines and chemokines. IL-6 levels significantly increased in the plasma of irAE patients compared to all 3 control groups, while IL-12p70 levels significantly increased in irAE patients compared to HC and ICI controls (Fig. 5A). IFN-γ, TNF-α, and IL-1β only showed a slight increase in irAE patients (Fig. 5B). Other cytokines, including IL-2, IL-4, IL-8 and IL-17A exhibited no clear differences among different groups (fig. S5A). Chemokines IP-10, CXCL11, CXCL9, ligands for CXCR3, were significantly elevated in the irAE patients compared to HC and RAC, but not ICI controls (Fig. 5C). Similar changes were observed on CCL20, a ligand for CCR6 (Fig. 5D). While CCL17, one of the ligands for CCR4, did not show any clear differences among the groups of patients (fig. S5B). CX3CL1 has been reported to be elevated in seropositive RA patients (*61*), and reduced level of CX3CL1 in patients was associated with clinical response to treatment with abatacept or infliximab (*62*). Although CX3CL1 level did not increase in seronegative RA patients in our cohort, it significantly increased in irAE patients compared to HC or RA, but not ICI controls (Fig. 5E). CCL2, a chemokine that plays a key role in attracting monocytes and T cells, also showed significant increases in irAE and ICI patients (Fig. 5F). Other chemokines for recruiting monocytes, NK cells, polymorphonuclear leukocytes, eosinophils and neutrophils including CCL3, CCL4 (fig. S5C), CXCL1, CXCL5, CCL11 and CXCL8 (fig. S5D) did not show significant changes in irAE patients, which suggests that T cell trafficking might be preferentially affected in IA irAE. Hence, plasma IL-6, and IL-12p70, concentrations were significantly increased in irAE patients. Because of the significantly enriched IFN-α response genes in irAE B cells, CD4^+^ T cells and CD8^+^ T cells, we concluded that type I IFN was also enhanced in irAE patients, consistent with previous literature (*18*).

**Fig. 5.**
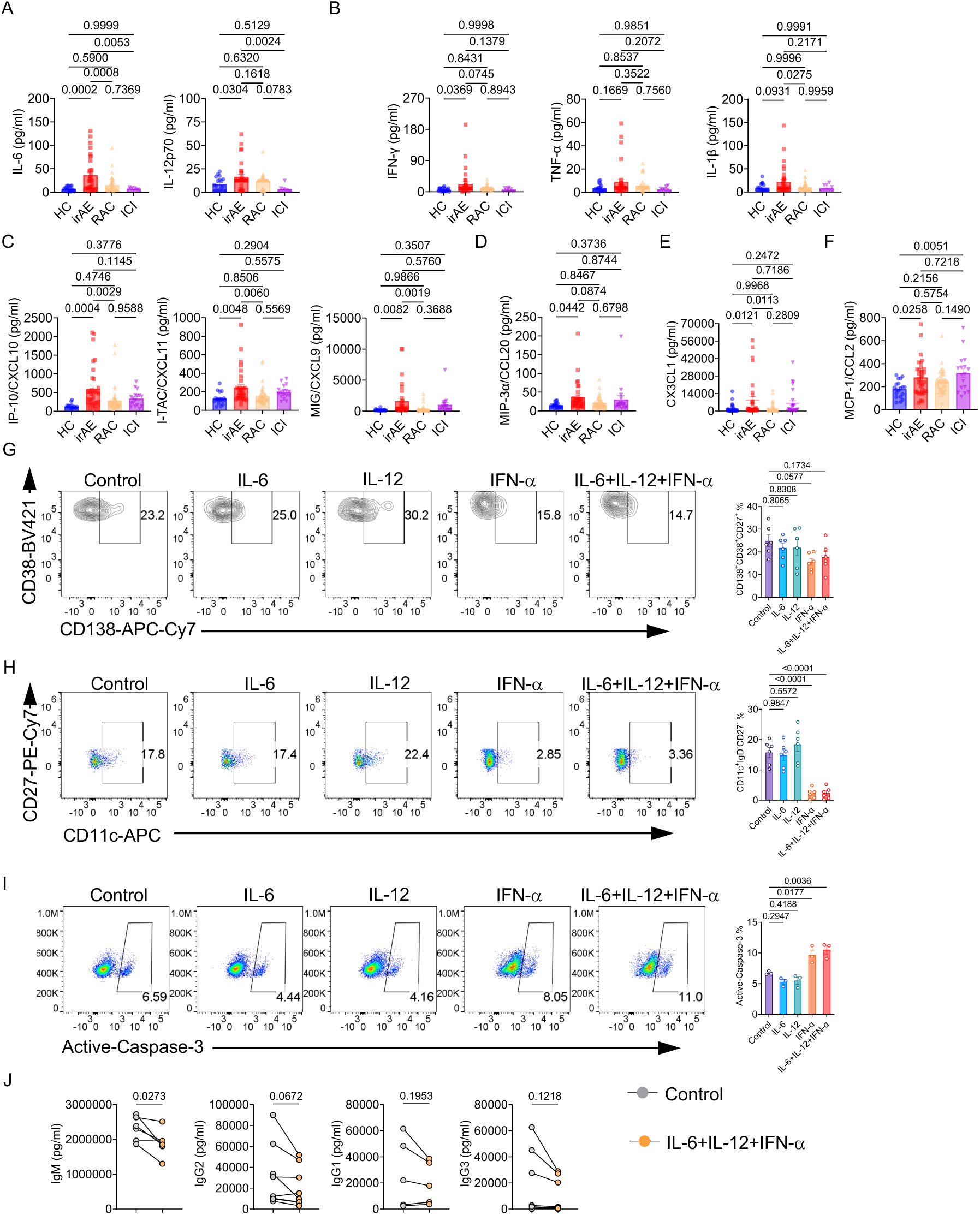
Inflammatory signatures enriched in irAE patients reduce antibody production (A-F) Beads based multiplex assays were used to measure plasma concentration of IL-6, and IL-12p70 (A), TNF-α, IFN-γ and IL-1β (B), HC (n = 19), irAE (n = 34), RAC (n = 45), ICI (n = 9). IP-10 (CXCL10), CXCL11, and CXCL9 (C, HC, n = 17; irAE, n = 33; RAC, n = 46; ICI, n = 17), CCL20 (D), CX3CL1 (E), and CCL2 (F). (G-J) Human naïve B cells were isolated and cultured with 0.5 μg/mL anti-human CD40, 2.5 μg/mL anti-human Ig (M+G+A), and 20 ng/mL rhIL-21 with 100 ng/mL IFN-α, 100 ng/mL IL-6, 100 ng/mL IL-12, control, or the combination of IFN-α, IL-6, and IL-12 for 7 days. Cells and culture supernatants were analyzed. (G) Representative flow plot of CD38 and CD138 expression on CD27^hi^CD38^hi^ ASCs. Right, a summary of the percentage of CD138^+^ ASCs, n = 6. (H) Expression of CD11c and CD27 on CD27^-^IgD^-^ ASCs. Right, percentage of CD11c^+^IgD^-^CD27^-^ B cells, n = 6. (I) Expression of active-caspase-3 in B cells. Right, percentage of active-caspase-3^+^ B cells from different groups, n = 3. (J) Different immunoglobulin isotype levels in the culture supernatants from G-H were measured by the multiplex assay, n = 6. Data in graphs represent mean ± SEM, Significance was tested by One-way ANOVA (A-I), and paired Student’s t-test (J).

Next, we investigated the impacts of IL-6, IL-12, and IFN-α on B cells. Human naïve B cells were cultured with IL-6, IL-12, IFN-α or their combination for 7 days. IFN-α and combined IL-6/IL-12/IFN-α significantly promoted the generation of CD38^hi^CD27^hi^ antibody-secreting cells and IgD^-^CD27^-^ B cells, while IL-6 and IL-12 had no obvious effects on the B cells (fig. S5E and S5F). However, CD138^+^ ASCs were slightly decreased upon IFN-α or a combination of IL-6, IL-12, and IFN-α stimulation (Fig. 5G). IFN-α and combined IL-6/IL-12/IFN-α significantly downregulated CD11c expression on IgD^-^CD27^-^ B cells (Fig. 5H). Furthermore, they significantly increased the percentage of active-caspase-3 in B cells (Fig. 5I) and 7-AAD^+^Annexin V^+^ B cells (fig. S5G), indicating increased cell death. Finally, combined IL-6/IL-12/IFN-α reduced IgM and IgG2 production (Fig. 5J). Therefore, IFN-α, or combined IL-6/IL-12/IFN-α increases B cell apoptosis and reduces production of IgM and IgG2. These data are broadly consistent with our observations that irAE patients exhibit largely normal B cell homeostasis and no clear increase in autoantibody production.

### Inflammatory cytokines IL-6, IL-12 and IFN-α promote both CD4^+^ and CD8^+^ T cell effector functions

We next explored the effects of IL-6, IL-12, and IFN-α on both CD4^+^ and CD8^+^ T cells. IL-6, IL-12, IFN-α or combined IL-6/IL-12/IFN-α had no effect on the proportion of CD45RA^-^CD8^+^ T cells on both day 1 and day 5 of the culture (fig. S6A) as well as the Ki67 expression in CD45RA^-^CD8^+^ T cells at day 5 (fig. S6B). However, IFN-α, and IL-6, IL-12 and IFN-α combination significantly increased the proportion of CD38^+^CD127^-^CD8^+^ T cells at both day 1 and day 5 (Fig. 6A) as well as CD69 expression (Fig. 6B, fig. S6C). Combined IL-6/IL-12/IFN-α also increased CD25 expression (Fig. 6C, fig. S6D). Meanwhile, IL-12 alone or combined IL-6/IL-12/IFN-α drastically increased the proportion of IFN-γ^+^granzyme B^+^ T cells (Fig. 6D) and Granzyme B^+^ T cells (fig. S6E). Along with the changes of effector or cytotoxicity, IL-12 alone or combined IL-6/IL-12/IFN-α increased the mitochondrial membrane potential (Fig. 6E), mitochondrial mass (Fig. 6F) and glucose uptake (Fig. 6G). Together, these data indicate that IL-12 alone or a combination of IL-12, IL-6, and IFN-α promotes cytotoxicity and metabolism of CD8^+^ T cells.

**Fig. 6.**
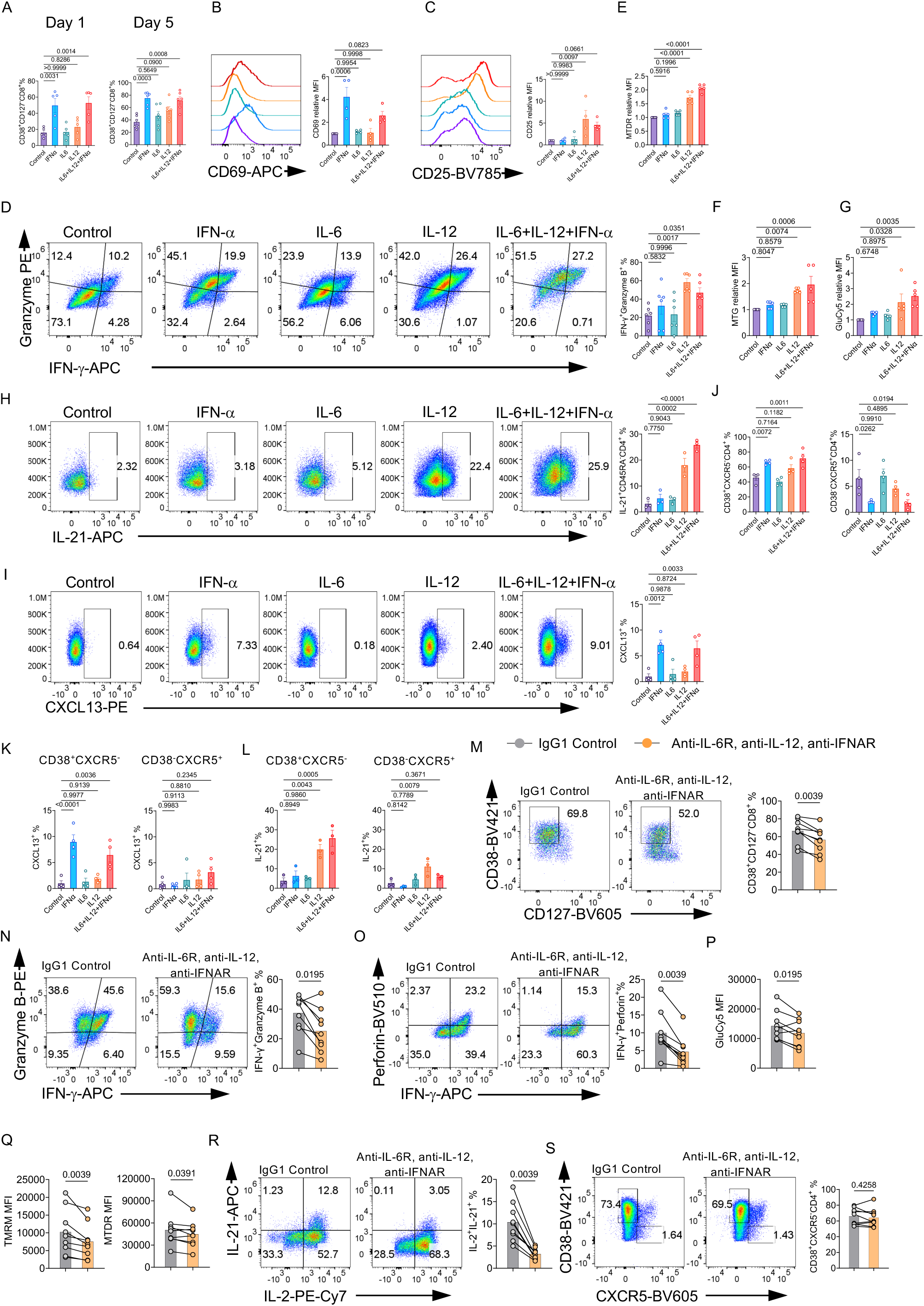
Inflammatory cytokines enriched in irAE patients promote both CD4^+^ and CD8^+^ T cell functions (A-G) CD8^+^ T cells were isolated from healthy donor PBMCs and cultured in the plate coated with 10 μg/mL anti-human CD3 and anti-human CD28 for indicated days in the presence of vehicle control (control), 100 ng/mL IFN-α, 100 ng/mL IL-6, 100 ng/mL IL-12, or the combination of IFN-α, IL-6 and IL-12. (A) Summaries of the percentage of CD38^+^CD127^-^CD8^+^ T cells at day 1 and day 5, n = 5. (B) CD69 expression was measured on day 5, and the mean fluorescence intensity (MFI) of CD69 was summarized on the right, n = 4. (C) CD25 expression was measured on day 5, and the MFI of CD25 was summarized on the right, n = 4. (D) CD8^+^ T cells activated for 5 days were restimulated with PMA/Ionomycin/Monensin for 5 h, expression of granzyme B and IFN-γ in CD8^+^ T cells were examined by flow cytometry. Right, percentage of granzyme B^+^IFN-γ^+^ CD8^+^ T cells, n = 5. (E-G) CD8^+^ T cells were cultured for 5 days. MFIs (all relative to those in control condition) of MTDR (E), MTG (F), and GluCy5 (G) were summarized, n = 5. (H-L) CD4^+^ T cells were isolated from the PBMCs of HC, cultured in the plate coated with 10 μg/mL anti-human CD3 and anti-human CD28 for indicated days in the presence of 100 ng/mL IFN-α, 100 ng/mL IL-6, 100 ng/mL IL-12, control or the combination of them. (H) CD4^+^ T cells activated for 5 days were restimulated with PMA/Ionomycin/Monensin for 5 h, and IL-21 level was examined in the CD45RA^-^CD4^+^ T cells, n =3. (I) Activated CD4^+^ T cells were restimulated with 10 μg/mL plate-coated anti-human CD3 and anti-human CD28 with monensin for 6 h, and expression of CXCL13 was measured in CD45RA^-^CD4^+^ T cells, n = 4. (J) CD38^+^CXCR5^-^ or CD38^-^CXCR5^+^ CD4^+^ T cells were examined at day 5, and the percentages were summarized, n = 4. (K) Percentage of CXCL13^+^ cells in CD38^+^CXCR5^-^ or CD38^-^CXCR5^+^ CD4^+^ T cells at day 5, n = 4. (L) Percentage of IL-21^+^ cells in CD38^+^CXCR5^-^ or CD38^-^CXCR5^+^ CD4^+^ T cells at day 5, n = 3. (M-S) PBMCs from the irAE patients were cultured in the plate coated with 10 μg/mL anti-CD3 and anti-CD28 in the presence of IgG1 isotype control or 50 μg/mL anti-human IL-6R, 50 μg/mL anti-human IL-12p40 and 50 μg/mL anti-human IFNAR1 for 3 days, n = 9. (M) Expression of CD38 and CD127 on CD8^+^ T cells. Right, percentage of CD38^+^CD127^-^CD8^+^ T cells. (N, O) CD8^+^ T cells activated for 5 days were restimulated with PMA/Ionomycin/Monensin for 5 h, (N) Expression of Granzyme B and IFN-γ. Right, percentage of granzyme B^+^IFN-γ^+^CD8^+^ T cells, (O) Expression of perforin and IFN-γ. Right, percentage of perforin^+^IFN-γ^+^CD8^+^ T cells. (P, Q) MFIs of GluCy5 (P), TMRM, and MTDR (Q) in CD8^+^ T cells were presented. (R, S) CD4^+^ T cells activated for 5 days were restimulated with PMA/Ionomycin/Monensin for 5 h, (R) Expression of IL-21 and IL-2. Right, percentage of IL-21^+^IL-2^+^ CD4^+^ T cells. (S) Expression of CD38 and CXCR5 on CD4^+^ T cells. Right, percentage of CD38^+^CXCR5^-^CD4^+^ T cells. Data in graphs represent mean ± SEM, Significance was tested by One-way ANOVA (A-L), and paired Student’s t-test (M-S).

CD4^+^ T cells not only promote humoral immune response but also facilitate CD8^+^ T cell function by secreting cytokines like IL-21 (*63–67*). IL-12, IL-6, IFN-α, alone, or in combination, did not affect the proportion of CD45RA^-^CD4^+^ cells (fig. S6F), or Ki67 expression on CD45RA^-^CD4^+^ T cells (fig. S6G), but IL-12 alone or combined IL-6/IL-12/IFN-α significantly prompted expression of IL-21 (Fig. 6H) and TNF-α (fig. S6H) in CD4^+^ T cells, while IFN-α alone or combined IL-6/IL-12/IFN-α raised the proportion of CXCL13^+^CD4^+^ T cells (Fig. 6I). Combined IL-6/IL-12/IFN-α also significantly upregulated IL-4 levels, while IFN-α alone increased the proportion of IL-2^+^CD45RA^-^CD4^+^ T cells (fig. S6I). IFN-α alone or combined IL-6/IL-12/IFN-α increased the proportion of CD38^+^CXCR5^-^CD4^+^ T cells, while reduced CD38^-^CXCR5^+^CD4^+^ T cells (Fig. 6J). Moreover, we further investigated possible differential effects of IL-6/IL-12/IFN-α on CD38^+^CXCR5^-^ vs CD38^-^CXCR5^+^ CD4^+^ T cells. IFN-α alone or the combination of IL-12, IL-6, and IFN-α elevated CXCL13 expression on CD38^+^CXCR5^-^CD4^+^ T cells, but not CD38^-^CXCR5^+^ CD4^+^ T cells (Fig. 6K). Importantly, CD38^+^CXCR5^-^CD4^+^ T cells expressed a higher level of IL-21 than CD38^-^CXCR5^+^ CD4^+^ T cells in response to IL-12 alone or a combination of IL-12, IL-6, and IFN-α stimulation (Fig. 6L). CD38^+^CXCR5^-^CD4^+^ T cells also had relatively higher levels of TNF-α (fig. S6J) and IL-4 (fig. S6K) compared to CD38^-^CXCR5^+^ CD4^+^ T cells, while they had comparable levels of IL-2 (fig. S6L). These data suggest that a combination of IL-12, IL-6, and IFN-α expands CD38^+^CXCR5^-^CD4^+^ T cells, and promotes the production of IL-21, IL-4, CXCL13, and TNF-α, which could impact other immune cells.

### Pharmacological Blocking IL-6, IL-12 and IFN-α attenuates elevated effector T cell phenotypes from irAE patients

Our above data supports a model that IL-6, IL-12, and IFN-α may contribute to some of the immunological aberrations found in irAE patients, which could be partially reversed by blocking these cytokines or their receptors. To test this hypothesis, we activated irAE PBMCs in the presence of combined anti-IL-6R, anti-IL-12, and anti-IFNAR. Combined IL-6/IL-12/IFN-α blockade significantly reduced the percentages of CD38^+^CD127^-^ (Fig. 6M), granzyme-B^+^IFN-γ^+^ (Fig. 6N), perforin^+^IFN-γ^+^ (Fig. 6O), and granzyme-B^+^perforin^+^ (fig. S6M) CD8^+^ T cells from irAE patients. Furthermore, they significantly downregulated glucose uptake (Fig. 6P), mitochondrial membrane potential (Fig. 6Q), but not mitochondrial mass (fig. S6N) of irAE CD8^+^ T cells. In terms of CD4^+^ T cells, combined IL-6/IL-12/IFN-α blockade significantly reduced IL-21 expression in irAE CD4^+^ T cells (Fig. 6R). However, IL-6/IL-12/IFN-α blockade did not affect the proportion of CD38^+^CXCR5^-^CD4^+^ T cells (Fig. 6S), nor did it significantly affect glucose uptake, mitochondrial membrane potential, mitochondrial mass of irAE CD4^+^ T cells (fig. S6, O-Q), suggesting a stronger impact of these cytokines on CD8^+^ T cells than CD4^+^ T cells. Together, these data indicate that elevated IL-6, IL-12, and IFN-α may contribute to the enhanced expression of some cytotoxicity molecules and metabolic activity in irAE CD8^+^ T cells.

## DISCUSSION

In this study, we aim to address two major questions, what are the immunological phenotypes that distinguish anti-PD-1/PD-L1 treated patients with IA irAE from those without irAE, and the relationship between IA irAE and RA. While previous studies have explored the immune cell phenotypes, our current study represents an in-depth immunological investigation on the largest cross-sectional IA irAE patient cohort with 3 different control groups so far, and thus provides robust information on this fast-growing disease owing to the increased usage of ICI agents. Our results demonstrate remarkable phenotypical changes in IA irAE CD4^+^ and CD8^+^ T cells. They include reduced Tfh-like CD4^+^ T cells, increased CXCR3^-^CCR6^-^ and reduced CXCR3^+^CCR6^+^ subsets in both CD4^+^ and CD8^+^ T cells, increased expression of effector and cytotoxic molecules, and heightened metabolic activity. Some of these are shared between IA irAE and ICI control, but many are unique in IA irAEs, including highly increased cytotoxic program in CD8^+^ T cells, reduced CXCR3 and CCR6 expression on CD4^+^ T cells, and increased metabolic activity in both CD4^+^ and CD8^+^ T cells. These results suggest that increased cytotoxic activity, but not simply increased expression of effector T cell markers, might be a prerequisite to IA irAE development. Such observation partly explains why irAE development is associated with improved tumor response to the ICI therapy (*68*) (Supplementary Table 1), i.e., those without irAE are more likely to have T cells with insufficient cytotoxic activity. This observation is consistent with the recent observation that CD38^hi^ cytotoxic CD8^+^ T cells are enriched in ICI-arthritis joints (*18*). Indeed, cytotoxic CD8^+^ T cells may directly contribute to RA pathogenesis (*69, 70*). The mechanisms through which reduced CXCR3 and CCR6 expression on CD4 T cells may contribute to IA irAE are unclear. Reduced CXCR3 and CCR6 on CD4 T cells are associated with increased ribosomal activity and protein translation, consistent with the increased metabolism in irAE T cells. Moreover, reduced CXCR3 is largely consistent with previous observation that CXCR3 ligands might promote T cell trafficking to joint tissues (*17*). Nevertheless, CXCR3^-^CCR6^-^ CD4^+^ T cells might be a biomarker for IA irAE. Furthermore, other T cell lineages, such as resident memory T cells, may contribute to IA irAE (*71, 72*).

In contrast to the drastic phenotypic changes in T cells, B cell compartment remains largely unaltered in IA irAE patients, whereas seronegative RA patients surprisingly exhibit more changes. RA control patients, but not IA irAE patients, have significantly increased CD4/CD8 ratio and frequency of CD11c^+^CD21^-^ atypical B cells. These atypical B cells accumulate during aging in healthy individuals, especially in females (*73, 74*). They are overrepresented in various systemic autoimmune disorders, such as systemic lupus erythematosus, Sjogren’s syndromes. Recent studies found an expansion of these cells in the synovium and peripheral blood of seropositive RA patients, indicating a possible role in RA pathogenesis (*73, 75, 76*). Here, for the first time we showed that these atypical B cells are also elevated in seronegative RA patients, consistent with the observation that seronegative RA patients have elevated levels of multiple autoantibodies (*57*). However, IA irAE patients do not exhibit elevated CD11c^+^CD21^-^ atypical B cells, nor increased levels of common autoantibodies. Moreover, PD-1 blockade does not consistently promote B cell activation or antibody production. Although a negative argument may require extraordinarily exhaustive proof, our data in its totality suggest that IA irAE is primarily associated with overactivation of T cell immunity but not B cell immunity. Hence, our study establishes IA irAE as an immunologically distinct subtype of arthritis and represents the first comprehensive immunological study of a unique T cell driven, and likely autoantibody-independent arthritis in human patients. Further analyses using larger sample size and study on the deposition of ICs and complement in joint tissues from IA-irAE patients are warranted in future investigations.

Another characteristic of IA irAE is the elevated levels of selective cytokines and chemokines, some of which (e.g., CCL2) are shared with ICI controls and most of which are absent in RA controls. These data suggest a complex inflammatory environment elicited by PD-1 inhibition and a relatively low inflammatory condition for seronegative RA. Several biologics have been suggested to treat IA irAEs, including tocilizumab (*77*), and TNF-α inhibitor (*17*), and type I IFN blockers (*18*). Our results indicate that IL-6, IL-12 and type I IFN likely play a role in promoting cytotoxic activity in IA irAE T cells. ICI treatment can promote IL-6 production, which contributes to irAE development (*78*). IL-12 is critical for successful anti-PD-1 therapy (*79*). Type I IFNs may have opposing roles in ICI therapy depending on the timing of its activation, i.e., promoting immunity early while suppressing immunity later on (*80*). Thus, while targeting combined IL-6/IL-12/IFN-α may benefit IA irAE, it carries the risk of compromising anti-tumor immunity. Thus, thoughtful consideration is needed to balance the management of irAE and maintenance of tumor control.

There are several limitations in our study. First, although our cohort is relatively large, more definitive answers on the autoantibody profile will require a much larger sample size and a genome wide autoantibody scan. Second, our study is a cross-sectional study. A longitudinal study will be needed to identify immunological phenotypes before and after ICI therapy, and before and after irAE onset. Lastly, all the cellular analyses were performed on PBMC samples. Paired joint samples will be needed to identify immune cells responsible for arthritis *in situ*.

## METHODS AND MATERIALS

### Study design

This study was approved by the institutional review board at Mayo Clinic (IRB protocol #21-009862), and patients providing written informed consent were eligible for this study.

Specifically, 4 groups of patients were recruited for this study, IA irAE (irAE) patients are cancer patients on PD-1 or PD-L1 inhibitors (pembrolizumab, nivolumab, cemiplimab, atezolimumab, durvalumab, or avelumab) therapy without pre-existing rheumatic diseases, who develop physician confirmed inflammatory arthritis; ICI-control (ICI) patients are cancer patients on PD-1 or PD-L1 inhibitors therapy without pre-existing rheumatic diseases, who have not developed any irAE; RA-control (RAC) patients are non-cancer patients with rheumatoid arthritis that match those with de novo rheumatic irAE; and heathy control (HC) are the sex- and age-matched healthy individuals.

### Mice

HuPD-1 and HuPD-L1 humanized mice were a gift from Dr. Haidong Dong at Mayo Clinic (*60*). Mice were bred and maintained in a specific pathogen-free facility in the Department of Comparative Medicine of the Mayo Clinic. The mice were euthanized by carbon dioxide according to the approved protocol. All animal protocols (A00003354-18-R23) were approved by the Institutional Animal Care and Use Committees (IACUC) of the Mayo Clinic Rochester.

### Samples collection and processing

Blood samples were collected into sodium heparin tubes, and plasma was collected after centrifuging at 4 °C, 1300 rpm for 10 mins. Peripheral blood mononuclear cells (PBMCs) were isolated from the peripheral blood using Ficoll-Paque gradient centrifugation. Specifically, 13 mL Ficoll-Paque™ PLUS density gradient media (GE Healthcare, Cat# 17-1440-02) were added to the bottom of 30 mL diluted peripheral blood, and centrifuged at 4 °C, 400 g for 25 min with the acceleration and deceleration at lower speed. The PBMC layer was collected and washed with wash buffer (1×PBS + 10 mM HEPES + 0.1% BSA + 2 mM EDTA), and PBMCs were aliquoted and cryopreserved in a liquid nitrogen tank.

### Flow cytometry

Cryopreserved PBMCs were thawed, rested, and washed. For surface staining, cells were stained in phosphate-buffered saline (PBS) containing 1% (w/v) bovine serum albumin (BSA) (FACS buffer) with indicated antibodies for 1 h on ice. Following antibodies have been used: anti-IgD-FITC (Biolegend, IA6-2, Cat# 348206), anti-CD24-PE (Biolegend, ML5, Cat# 311106), anti-IgM-Percp-Cy5.5 (Biolegend, MHM-88, Cat# 314512), anti-CD21-APC (BD Pharmingen, B-ly4, Cat# 561357), anti-CD11c-redFluor™ 710 (CyTEK, 3.9, Cat# 80-0116-T025), anti-CD138-APC-Cy7 (Biolegend, MI15, Cat# 356528), anti-CD27-PE-Cy7 (Biolegend, M-T271, Cat# 356412), anti-CD38-BV421 (Biolegend, HIT2, Cat# 303525), anti-CD3-Super Bright 600 (Invitrogen, OKT3, Cat# 63-0037-42), anti-CD19-BV650 (Biolegend, HIB19, Cat# 302238), anti-CD14-BV785 (Biolegend, M5E2, Cat# 301840), CXCR5-BV605 (Biolegend, J252D4 Cat# 356930), anti-CXCR5-AF647 (Biolegend, J252D4, Cat# 356906), anti-CD45RA-APC-Cy7 (CyTEK, HI100, Cat# 25-0458-T100), anti-CD28-BUV395 (BD Horizon, CD28.2, Cat# 569160), anti-CD38 (Biolegend, HIT2, Cat# 303504), anti-CCR6-PE (Biolegend, G034E3, Cat# 353410), anti-CX3CR1-Percp-Cy5.5 (Biolegend, 2A9-1, Cat# 341614), anti-CD4-AF700 (Biolegend, RPA-T4, Cat# 300526), anti-CCR7-PE-Cy7 (Biolegend, G043H7, Cat# 353226), anti-CD8-BV510 (Biolegend, RPA-T8, Cat# 301048), anti-CD127-BV605 (Biolegend, A019D5, Cat# 351334), anti-CD3-BV650 (Biolegend, OKT3, Cat# 317324), anti-PD-1-BV711(Biolegend, EH12.2H7, Cat# 329928), anti-CD25-BV785 (Biolegend, BC96, Cat# 302636), anti-CXCR3-BV421 (Biolegend, G025H7, 353716), anti-CD69 (Biolegend, FN50, Cat# 310904). For the cytokine intracellular staining, PBMCs were stimulated with phorbol 12-myristate 13-acetate, ionomycin and GolgiSTOP in a complete RPMI1640 medium (RPMI1640 + 10% (vol/vol) FBS + 1% penicillin-streptomycin-L-Glutamine) for 5 h. Cells were stained for surface molecules, fixed with BD Fixation/Permeabilization solution (BD CytoFix/CytoPerm^TM^, Cat# 51-2090KZ) on ice for 20 min, permeabilized with BD Perm/Wash^TM^ buffer (BD Perm/ Wash^TM^ solution, Cat# 51-2091KZ), and stained with the following antibodies: anti-TNF-α-AF488 (Biolegend, MAb11, Cat# 502915), anti-Granzyme B-PE (Invitrogen, GB11, Cat#12-8899-41), anti-IL-2-PE-Cy7 (Biolegend, MQ1-17H12, Cat# 500326), anti-IFN-g-APC (Biolegend, 4S.B3, Cat# 502512), anti-IL-21-APC (Biolegend, 3A3-N2, Cat#513008), anti-IL-4-PE (Biolegend, MP4-25D2, Cat# 500810), anti-perforin-BV510 (Biolegend, dG9, Cat# 308120). For CXCL13 staining, cells were stimulated with 10 μg/mL coated anti-CD3, anti-CD28, and Monensin Solution for 5 h. Cells were stained with surface molecules, fixed with BD Fixation/Permeabilization solution, permeabilized with BD Perm/WashTM buffer, and stained with anti-CXCL13-PE (R&D, 53602, Cat#IC8012P). For the transcriptional factor staining, cells were stained with surface molecules, fixed at Room temperature with True-Nuclear Fix buffer for 30 mins, permeabilized with True-Nuclear perm buffer, and stained with anti-Ki67-BV421 (BD Horizon, B56, Cat# 562899). Cell viability was examined by Fixable viability dye (Tonbo Bioscience) or 7-AAD (Thermo Fisher).

For the staining of metabolic indicators, cells were washed with PBS, stained with viability dye and the indicated metabolic indicators, including 20 nM MitoTracker Deep Red (ThermoFisher Scientific), 20 nM MitoTracker Green (ThermoFisher Scientific), 100 nM tetramethylrhodamine methyl ester (TMRM, ThermoFisher Scientific), 500 nM CellROX or 1 μM GluCy5 in HBSS at 37 °C for 20 min, following by the surface molecules staining. Flow cytometry was performed on a BD Fortessa X-20, LSR II instrument, or Attune NxT system (Life Technologies). Data were then analyzed by FlowJo software (Tree Star).

### Cytokine and immunoglobulin measurement

For the cytokine, chemokine, and immunoglobulin levels in the human plasma, LEGENDplex™ kits from Biolegend were used according to the manufacturer’s instructions. Specifically, plasma samples were diluted 2-fold or 100,000-fold (For immunoglobulin) using assay buffer, and the following kits were used in this study: Human Essential Immune Response Panel (Cat# 740930), Human Proinflammatory Chemokine Panel 1(Cat# 740985), Human B Cell Activator Panel (Cat# 740535), Human Immunoglobulin Isotyping Panel (Cat# 740640). The data were acquired on the Attune NxT system (Life Technologies) and analyzed using cloud-based software from Qognit. For CXCL13, IL-21, and CX3CL1 measurements, the following kits were used: Human CXCL13/BLC/BCA-1 Quantikine ELISA (R&D system, Cat# DCX130), Human IL-21 DuoSet ELISA (R&D system, Cat# DY8879-05), Human CX3CL1/Fractalkine DuoSet ELISA (R&D system, DY365), and all steps were performed according to the manufacturer’s instructions. The plates were read at 450 nM with the wavelength correction of 570 nM.

### Human naïve B cell pulsed with pembrolizumab

Cryopreserved PBMCs were thawed, rested, and washed. Human naïve B cells were isolated from PBMCs using the EasySep™ Human Naïve B Cell Isolation Kit (Stemcell Technologies, Cat# 17254), and only the samples with the percentage of IgD^+^CD27^-^ over 95% were used for the culture experiment. Specifically, three conditions were adopted(*14, 59, 81, 82*) and used in this study. Condition 1: 2.5 μg/mL anti-human Ig (M + G + A) (Jackson Immunoresearch, Cat# 109-006-064), 2.5 μg/mL CpG ODN (Invivogen, Cat# tlrl-2006-1), 10 μg/mL anti-human CD40 (BioXcell, Cat# BE0189), 20 ng/mL rhIL-21 (Peprotech, Cat# 200-21-50UG), 10 ng/mL rhIL-4 (Biolegend, Cat# 574004), 10 ng/mL rhIL-2 (Peprotech, Cat#200-02-250UG). Condition 2: 0.5 μg/mL anti-human CD40, 20 ng/mL rhIL-21, 2.5 μg/mL anti-Ig (M + G + A). Condition 3: 0.5 μg/mL R848 (Invivogen, Cat# tlrl-r848-1), 20 ng/mL rhBAFF (Biolegend, Cat# 7449534), 10 ng/mL rhIL-2, 10 μg/mL anti-human CD40, 2.5 μg/mL anti-human Ig (M + G + A),10 ng/mL rhIL-21, 20 ng/mL rhIFN-γ (Biolegend, Cat# 570204). 100 μg/mL pembrolizumab (Keytruda) or Isotype control IgG4 (BioXcell, Cat# CP147) was added into the culture media 2 days after stimulation, and B cells were further cultured for another 5 days. Surface molecules, including IgD, CD86, CD138, CD27, CD38, CD11c, and PD-1, were examined by flow cytometry, and the supernatant was used to measure immunoglobulin levels (Legendplex Human immunoglobulin isotyping panel, Biolgend).

### IL-6, IL-12 and IFN-α stimulation on human naïve B cells

Naïve B cells were isolated from healthy donor PBMCs using EasySep™ Human Naïve B Cell Isolation Kit (Stemcell Technologies, Cat# 17254). B cells were stimulated with 0.5 μg/mL anti-human CD40, 2.5 μg/mL anti-human Ig (M+G+A), 20 ng/mL rhIL-21 with or without 100 ng/ml rhIL-6 (Biolegend, Cat#570806), 100 ng/ml rhIL-12 (Biolegend. Cat# 573004), 100 ng/ml rhIFN-α2 (Biolegend, Cat# 592704) or the combination of 100 ng/ml rhIL-6, 100 ng/ml rhIL-12, and 100 ng/ml rhIFN-α for 7 days. Culture supernatants were collected, and cells were analyzed by flow cytometry.

### Human T cell culture

CD4^+^ T cells or CD8^+^ T cells were isolated from PBMCs using the EasySep™ Human CD4^+^ T Cell Enrichment Kit (Stemcell Technologies, Cat# 19052), EasySep™ Human CD8^+^ T Cell Isolation Kit (Stemcell Technologies, Cat# 17953), respectively. Only the samples with purity over 95% were used in the experiment. 0.5 million isolated CD4^+^ T cells or CD8^+^ T cells were stimulated with 10 μg/mL plate-coated anti-CD3 (Bio X cell, Cat# BE0001-2) and anti-CD28 (Bio X cell, Cat# BE0291), 100 ng/mL rhIL-6, 100 ng/mL rhIFN-α2, 100 ng/mL rhIL-12, or the combination of rhIL-6, rhIFN-α2 and rhIL-12 for 5 days. Surface molecules CD38, CXCR5, CD127, CD45RA, CD69, and CD25 were measured on the cultured T cells, and Ki67 and cytokine levels were also evaluated on the T cells.

### Humanized PD-1 mouse B cell culture

Mouse B cells were isolated from splenocytes using the EasySep Mouse B Cell Isolation Kit (Stemcell Technologies, catalog no. 19854). B cells were labeled with CTV (Celltraceviolet, Invitrogen) and cultured in RPMI1640 medium supplemented with 10% (vol/vol) FBS and 1% penicillin-streptomycin. Three conditions were used in this study. Condition 1: 3 μg/mL LPS (Sigma Aldrich, Cat# L2880-25MG), 10 ng/mL recombinant mouse IL-4 (R&D, Cat# 404-ML-100/CF) plus 20 ng/mL recombinant human BAFF (Biolegend, Cat# 7449534). Condition 2: 5 μg/mL Anti-mouse IgM (Jackson Immunoresearch, Cat# 115-006-075), 2.5 μg/mL CpG ODN, 100 ng/mL rmIL-21 (Biolegend, Cat# 570502), 20 ng/mL rmIL-4, 100 ng/mL rhIL-2. Condition 3 (*59*): 1 μg/mL R848, 1 μg/mL anti-mouse CD40 (BioXcell, Cat# BE0016-2), 1 μg/mL anti-mouse IgM, 100 ng/mL rmIL-21, 10 ng/mL rmIFN-γ (Peprotech, Cat# 315-05 20 ug). B cells were cultured for 3 days, 100 μg/mL pembrolizumab or Isotype control IgG4 was added on Day 1.

### Puromycin incorporation

Puromycin incorporation assay was adopted (*83*). Briefly, cells were treated with control, 100 mM 2-Deoxy-D-Glucose (DG), 1μM Oligomycin (Oligo), or a sequential combination of the drugs at the final concentrations for 30-45 mins, 10 μg/mL puromycin (Invitrogen) was incubated with cells for 20 min. After puromycin treatment, cells were washed in cold PBS and stained with a combination of Fc receptors blockade and cell viability dye, then primary conjugated antibodies against surface markers for 30 min on ice in FACS buffer. After washing, cells were fixed and permeabilized using True-Nuclear™ Transcription Factor Buffer Set (Biolegend, Cat# 424401) following manufacturer instructions. Intracellular puromycin staining was performed with anti-puromycin-AF647 (Sigma Aldrich, 12D10, Cat# MABE343-AF647) by incubating cells for 1 h on ice. The MFI was used to calculate mitochondrial and glucose metabolism. Briefly, Mitochondria metabolism = (Con– DGO) – (2-DG-DGO), Glucose metabolism = (Con– DGO) – (2-DG-O), DMSO control (Con), DGO (2-DG + Oligomycin), O (Oligomycin).

### Flow cytometric cell sorting for bulk RNA-seq

Human PBMCs were thawed and rested in a complete medium for 1 h; CD4^+^ or CD8^+^ T cells were enriched using EasySep™ Human CD4^+^ T Cell Isolation Kit and EasySep™ Human CD8^+^ T Cell Isolation Kit, respectively. Enriched CD4^+^ T cells were stained with PE-anti-human CCR6, 7-AAD, PE-Cy7-anti-human CD4 and BV421-anti-human CXCR3, while enriched CD8^+^ T cells were stained with PE-anti-human CCR6, 7-AAD, BV510-anti-human CD8 and BV421-anti-human CXCR3. CXCR3^-^CCR6^-^, CXCR3^+^CCR6^-^, CXCR3^-^CCR6^+^, or CXCR3^+^CCR6^+^ cells were sorted from CD4^+^ or CD8^+^ T cells using BD FACSAria™ III Cell Sorter. The sorted cells were lysed, and total RNA was extracted using Quick-RNA^TM^ Micro Prep (Zymoresearch, Cat# R1051). After quality control, high-quality total RNA was used to generate the RNA sequencing library using the DNBSEQ platform at Innomics. Reads with low quality, containing the adapter (adapter pollution), or with high levels of N base were removed to generate clean data. HISAT (V2.2.1) was used to align the clean reads to the Homo_sapiens reference genome (Homo_sapiens_9606.NCBI.GCF_000001405.39_GRCh38.p13.v2201) was used to align the clean reads to the reference genes. DEG analysis was carried out using DESeq2, and genes with log2FC > 0 and false discovery rate < 0.05 were considered for gene cluster analysis.

### Anti-IL-6R, anti-IL-12 and anti-IFNAR treatment on PBMCs from irAE patients

PBMCs from irAE patients were thawed and rested in a complete medium for 1 h, then activated with 10 μg/mL plate-coated anti-human CD3 and anti-human CD28 with IgG1 isotype control (BioXcell, Cat# CP174) or a combination of 50 μg/mL anti-human IL-6R (BioXcell, Cat# SIM0014), 50 μg/mL anti-human IL-12p40 (BioXcell, Cat# SIM0020) and 50 μg/mL anti-human IFNAR1 (BioXcell, Cat# SIM0022) for 3 days. Cells were treated with PMA, ionomycin, and monensin for 5 hours; surface molecules were stained, followed by intracellular cytokine staining using BD Cytofix/Cytoperm™ Fixation/Permeabilization Kit according to the manufacturer’s instruction.

### Sample processing for scRNAseq and scTCRseq

Cryopreserved PBMC samples were thawed and recovered in the RPMI1640 medium supplemented with 10% (vol/vol) FBS and 1% penicillin-streptomycin for 2 hours at 37 °C in 5% CO_2_. PBMCs were filtered with 40 μm nylon mesh and washed with PBS supplemented with 0.04% (vol/vol) BSA. The cell concentration was adjusted to 5 × 10^5^ /ml, cell counts and viabilities were determined using trypan blue exclusion and counted on a Countess II FL automated cell counter (Life Technologies), and only the PBMCs with viability over 90% were sent for sequencing. Reagents, reaction master mixes, reaction volumes, cycling numbers, cycling conditions, and clean-up steps were performed according to 10X Genomics’ guidelines. cDNA was allocated for preparation of a gene expression library (Chromium Single Cell 5’ Library Construction Kit; Cat#1000020) or TCR enrichment/library preparation (Chromium Single Cell V(D)J Enrichment Kit, Human T cell; Cat# 1000005). Sequencing was performed on an Illumina NovaSeq S4 platform.

### scRNAseq + scTCRseq data processing and analysis

FASTQ files generated from the Gene Expression (GEX) and Feature Barcode (TCR) libraries were processed using the 10x Genomics Cell Ranger multi pipeline (v7.0.0) to create expression matrices for downstream analysis. Integrated single-cell analysis of gene expression and T cell receptor data (scGEXseq + scTCRseq) was performed using the Immunopipe package (v1.4.0) (*84*). Cell type annotation was conducted using the Azimuth package with the Human PBMC reference dataset (*30*). To refine cell type classifications, we manually reviewed cluster-specific gene markers identified by Immunopipe and further annotated subpopulations of CD8 T cells. Gene set enrichment analysis (GSEA) was performed using the clusterProfiler package (v4.10.1) in R (*85*). Additionally, ANOVA test was applied to compare gene expression scores across patient groups for selected genes involved in the CD8 T cell cytotoxicity pathway (*86*).

### Autoantigen array

Autoantibody profiling was performed using an OmicsArray™ antigen microarray platform (Genecopoeia, Rockville, MD). This platform has the capacity to display large number of antigens on a 3D surface of a biochip and thereby serves as a multiplex screening method for the determination of autoantibody specificities. In this study, we used systemic autoimmune-associated Antigen Array (cat# PA001) panel that contains 120 autoantigens and 8 internal controls. A while-chip citrullination process was performed to citrullinate 120 autoantigens using a peptidyl arginine deiminase (PAD) cocktail that contained a mixture of 4 PAD isoforms (PAD1, PAD2, PAD3, PAD4) (*87, 88*). The non-citrullinated (non-cit) and citrullinated (cit) antigen arrays were run in parallel on a cohort of 125 human serum samples for profiling IgG and IgM antibody reactivities. Briefly, 2 <u>m</u>l serum samples were pre-treated with 1 unit of DNAse-I to remove free DNA, then diluted at 1:100 with PBST and hybridized onto antigen arrays. The antibodies binding with the antigens on the array were detected with Cy3-conjugated anti-human IgG and Cy5-conjugated anti-human IgM (1:2000, Jackson ImmunoResearch Laboratories). The fluorescent images were acquired with a Genepix 4000B scanner (Molecular Devices, San Jose, CA) and the signals were converted to signal intensity values using GenePix 7.0 software (Molecular Devices). Background was subtracted, and the net signal was normalized to internal controls for IgG and IgM, respectively. The final value for each autoantibody was expressed as normalized net signal intensity (NSI-nor).

All autoantigens from the arrays were log10-transformed, then standardized by subtracting the overall mean and dividing by the overall standard deviation. Association between each autoantigen and irAE versus healthy-, RA-, and ICI-controls (as well as RA vs healthy controls) were assessed using logistic regression adjusted for age and sex with indicator for irAE as the outcome and log10-transformed, standardized autoantigen value on the right-hand side.

Additionally, all comparisons with healthy controls were limited to females less than 64 years of age to match the makeup of the healthy controls for comparability. Results from each of the comparisons were summarized as volcano plots, with -log10 p-values plotted on the y-axis against the effect estimates on the x-axis. In addition, effect estimates were summarized as heatmaps. In all analyses, p-values less than 0.05 were considered statistically significant. Analysis was done using SAS version 9.4M8 (SAS Institute, Cary, NC, USA) and R version 4.2.2 (R Foundation for statistical computing, Vienna, Austria).

## Data Availability

All data produced in the present study are available upon reasonable request to the authors

## ACKNOWLEDGEMENT

We would like to thank Jane Jaquith, Kathleen McCarthy-Fruin, and Annie Streicher for helping with patient recruitment. We thank all the patients for donating their samples. We would like to thank Dr. Quanzhen Li and his team at Genecopoeia, Inc. for autoantigen array and data analysis. Additionally, we are grateful for the insightful discussions with Drs. Cornelia Weyand, Jörg Goronzy, and Ines Sturmlechner.

## Author contributions

X.Z. U.T. and H.Z. designed the project. U.T. and H.Z. administered, supervised, and acquired funding for the project. X.Z., Y.Y., Y.L., P.W., Y.L., H.D., and H.Z. contributed to the methodology. Y.Y., P.W., Y.L., and C.M. performed bioinformatics analysis. X.Z. performed most of the experiments. Y.L. managed the animal colony. B.E.S., S.N.M. and J.M.D. consented and recruited the patients. S.C. and U.T. reviewed the clinical records. H.D. provided the humanized mouse model and antibodies. H.E.L., A.C.H. and C.S.C. performed statistical analysis. X.Z. and H.Z. wrote the original draft, which was revised and edited by X.Z., C.S.C., S.N.M., J.M.D., H.D., U.T., and H.Z.

## Data and material availability

All data associated with this study are present in the paper or Supplementary Materials. Sequencing data is available at GEO (accession number xxx).

---

**Fig. S1.**
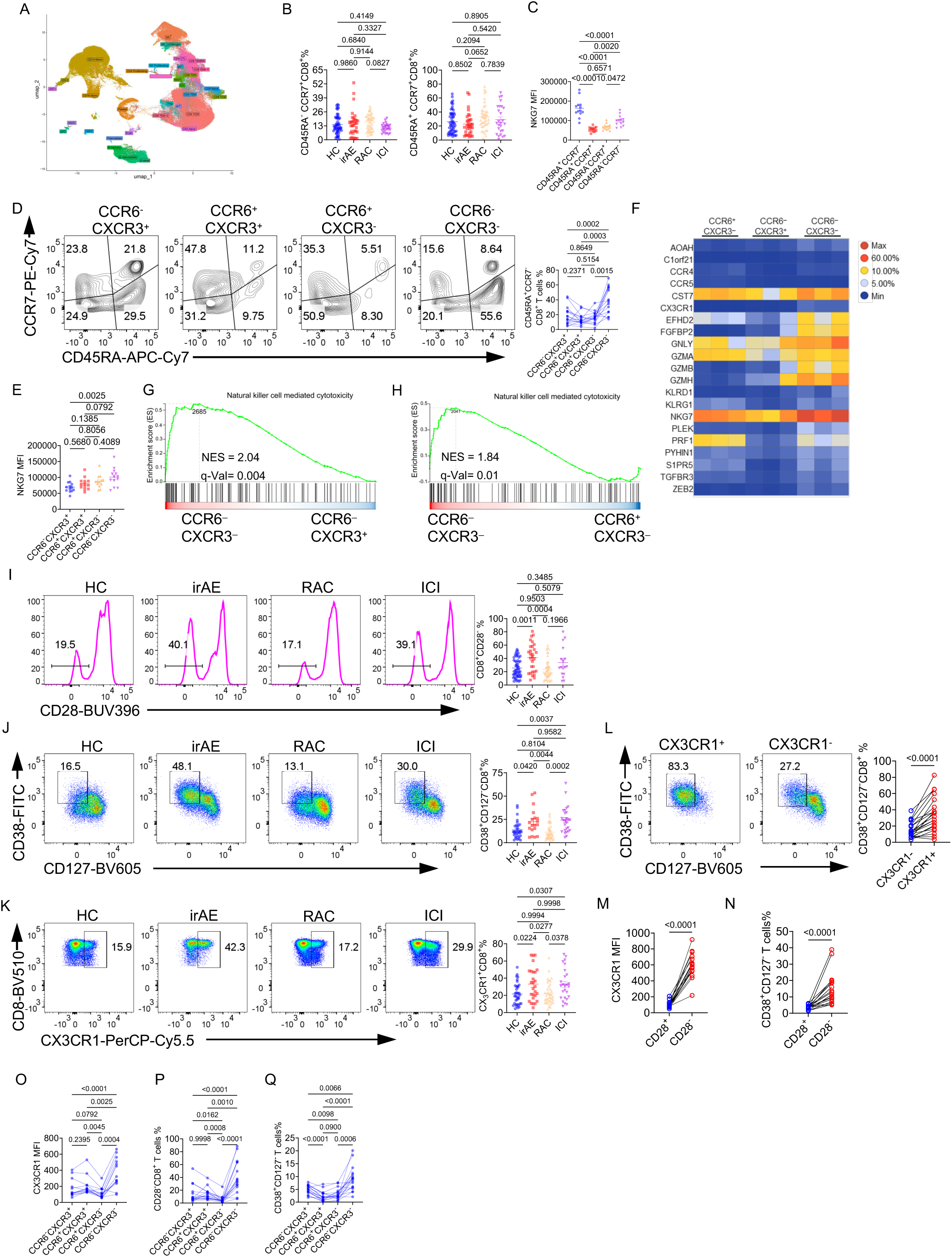
CD8^+^ T cells from irAE patients exhibit higher effector functions (A) UMAP and the cluster annotation of major immune cells in the PBMCs from HC (healthy control), ICI (ICI control), RAC (RA control), and irAE (IA irAE), and a total of around 220,000 cells were sequenced. (B) Summaries of the percentage of CD45RA^-^CCR7^+^CD8^+^ central memory T cells and CD45RA^+^CCR7^+^CD8^+^ naïve T cells, HC (n = 53), irAE (n = 29), RAC (n = 41), ICI (n = 17). (C) Summaries of the NKG7 mean fluorescence intensity (MFI) on CD45RA^+^CCR7^-^, CD45RA^+^CCR7^+^, CD45RA^-^CCR7^+^ or CD45RA^-^CCR7^-^ CD8^+^ T cells, n = 13. (D) Expression of CD45RA and CCR7 on CXCR3^+^CCR6^-^, CXCR3^+^CCR6^+^, CXCR3^-^CCR6^+^ or CXCR3^-^CCR6^-^ CD8^+^ T cells. Right, a summary of the percentage of CD45RA^+^CCR7^-^ (TEMRA) in different populations, n = 16. (E) Summary of the NKG7 MFI on CXCR3^+^CCR6^-^, CXCR3^+^CCR6^+^, CXCR3^-^CCR6^+^ or CXCR3^-^CCR6^-^ CD8^+^ T, n = 13. (F-H) CXCR3^+^CCR6^-^CD8^+^, CXCR3^-^CCR6^+^CD8^+^ or CXCR3^-^CCR6^-^CD8^+^ cells were sorted from healthy donors, and subjected to the bulk-RNA sequencing. (F) Heatmap for TEMRA signature genes expression. (G) Enrichment of Natural killer cell-mediated cytotoxicity between CXCR3^-^CCR6^-^CD8^+^ T cells and CXCR3^+^CCR6^-^CD8^+^ T cells. (H) Enrichment of Natural killer cell-mediated cytotoxicity between CXCR3^-^CCR6^-^CD8^+^ T cells and CXCR3^-^CCR6^+^CD8^+^ T cells. (I) Expression of CD28 on CD8^+^ T cells. Right, percentage of CD28^-^CD8^+^ T cells, HC (n = 53), irAE (n = 29), RAC (n = 41), ICI (n = 17). (J) Expression of CD38 and CD127 on CD8^+^ T cells. Right, percentage of CD38^+^CD127^-^CD8^+^ T cells, HC (n = 31), irAE (n = 16), RAC (n = 31), ICI (n = 15). (K) Expression of CX3CR1 on CD8^+^ T cells. Right, percentage of CX3CR1^+^CD8^+^ T cells, HC (n = 35), irAE (n = 24), RAC (n = 37), ICI (n = 17). (L) Expression of CD38 and CD127 on CX3CR1^+^CD8^+^ T cells or CX3CR1^-^CD8^+^ T cells. Right, percentage of CD38^+^CD127^-^ on CX3CR1^+^CD8^+^ T cells or CX3CR1^-^CD8^+^ T cells, n = 21. (M) Summary of the CX3CR1 MFI on CD28^+^ or CD28^-^ CD8^+^ T cells, n = 17. (N) Summary of the percentage of CD38^+^CD127^-^ T cells on CD28^+^ or CD28^-^ CD8^+^ T cells, n = 17. (O-Q) Summary of the CX3CR1 MFI (O), the percentage of CD28^-^ T cells (P), the percentage of CD38^+^CD127^-^ T cells (Q) on CXCR3^+^CCR6^-^, CXCR3^+^CCR6^+^, CXCR3^-^CCR6^+^ or CXCR3^-^CCR6^-^ CD8^+^ T cells, n = 16. Data in graphs represent mean ± SEM, Significance was tested by One-way ANOVA (A-K, O-Q), and paired Student’s t-test (L-N).

**Fig. S2.**
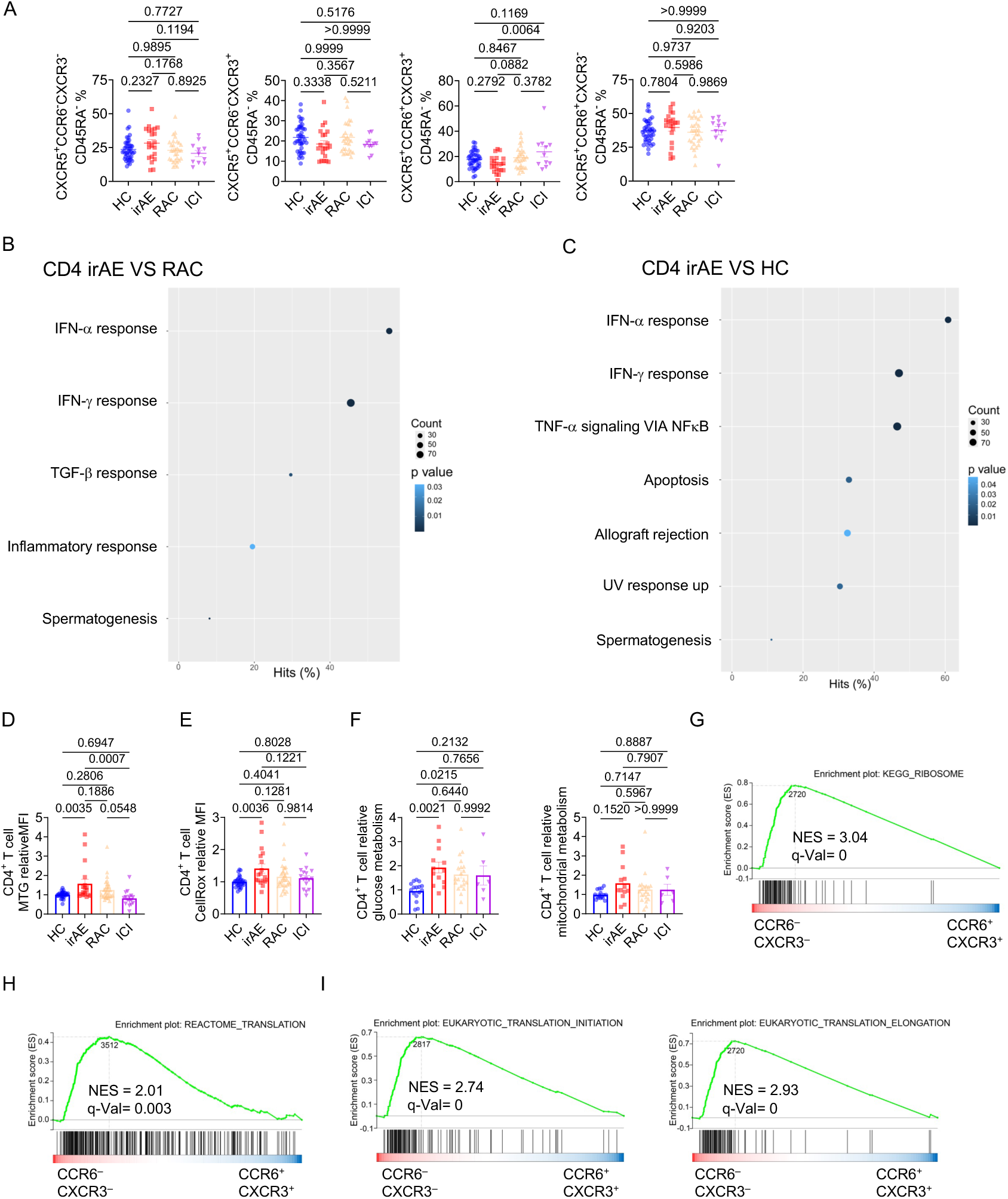
Distinctive CD4^+^ T cell phenotypes in irAE patients (A) Summaries of the percentage of CXCR3^-^CCR6^-^, CXCR3^+^CCR6^-^, CXCR3^+^CCR6^+^, CXCR3^-^ CCR6^+^ on CXCR5^+^CD45RA^-^CD4^+^ T cells, HC (n = 42), irAE (n = 21), RAC (n = 30), ICI (n = 11). (B) Significantly enriched pathways on CD4^+^ T cells from irAE vs RAC comparison. (C) Significantly enriched pathways on CD4^+^ T cells from irAE vs HC comparison. (D and E) PBMCs from different groups were stimulated with 10 μg/mL plate-coated anti-human CD3 and anti-human CD28 for 5 days, Mean fluorescence intensities (MFI) of MTG (D) and CellRox (E) in CD4^+^ T cells were presented, HC (n = 31), irAE (n = 18), RAC (n = 32), ICI (n = 16). (F) The puromycin incorporation assay was performed on fresh CD4^+^ T cells. Mitochondrial metabolism and glucose metabolism were calculated, HC (n = 16), irAE (n = 13), RAC (n = 20), ICI (n = 6). (G-I) CXCR3^-^CCR6^-^, CXCR3^+^CCR6^-^, CXCR3^+^CCR6^+^, CXCR3^-^CCR6^+^ cells were sorted from CD4^+^ T cells, and subjected to the bulk-RNA seq analysis. Enrichments of KEGG_Ribosome (G), Reactome Translation (H), and Eukaryotic_Translation_Initiation and Eukaryotic_Translation_Elongation (I) were evaluated between CXCR3^-^CCR6^-^ and CXCR3^+^CCR6^+^ CD4^+^ T cells. Data in graphs represent mean ± SEM, Significance was tested by One-way ANOVA.

**Fig. S3.**
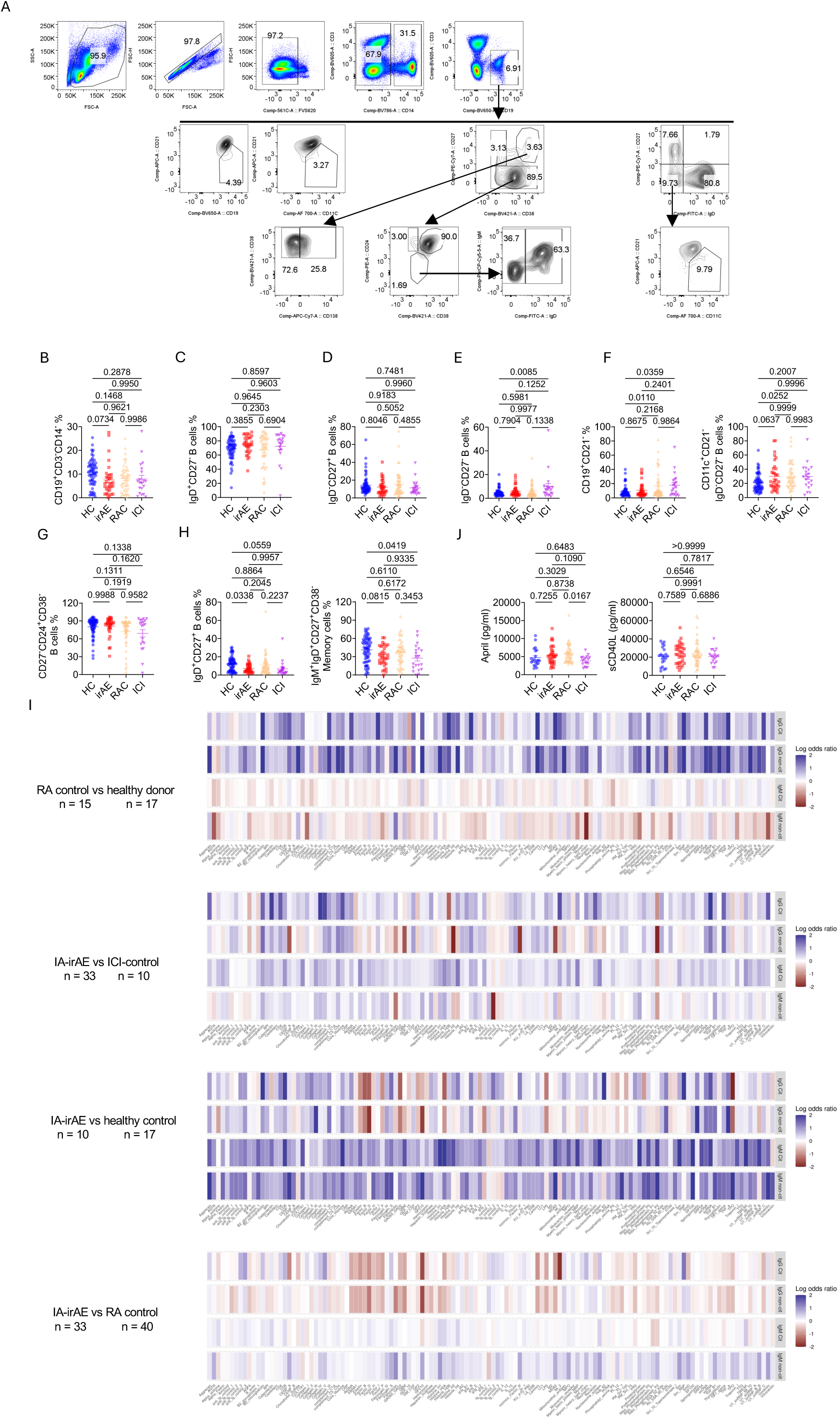
Humoral immunity is largely intact in irAE patients (A) Gating strategy for different B cell subsets in PBMCs was adopted from previous literature (*51*). (B) Summary of the percentage of CD19^+^CD3^-^CD14^-^ B cells in PBMCs, HC (n = 63), irAE (n = 33), RAC (n = 46), ICI (n = 20). Summaries of the percentage of IgD^+^CD27^-^ B cells(C), IgD^-^ CD27^+^ B cells (D), IgD^-^CD27^-^ double negative B cells (E), CD19^+^CD21^-^ B cells (F), CD27^-^ CD24^+^CD38^+^ transitional B cells (G), IgD^+^CD27^+^ non-switched memory B cells (H), and IgM^+^IgD^+^CD27^+^CD38^-^ non-switched memory B cells (H). (I) Heatmap of the autoantigen levels between different groups. (J) Plasma concentrations of April and sCD40L. Data in graphs represent mean ± SEM, Significance was tested by One-way ANOVA.

**Fig. S4.**
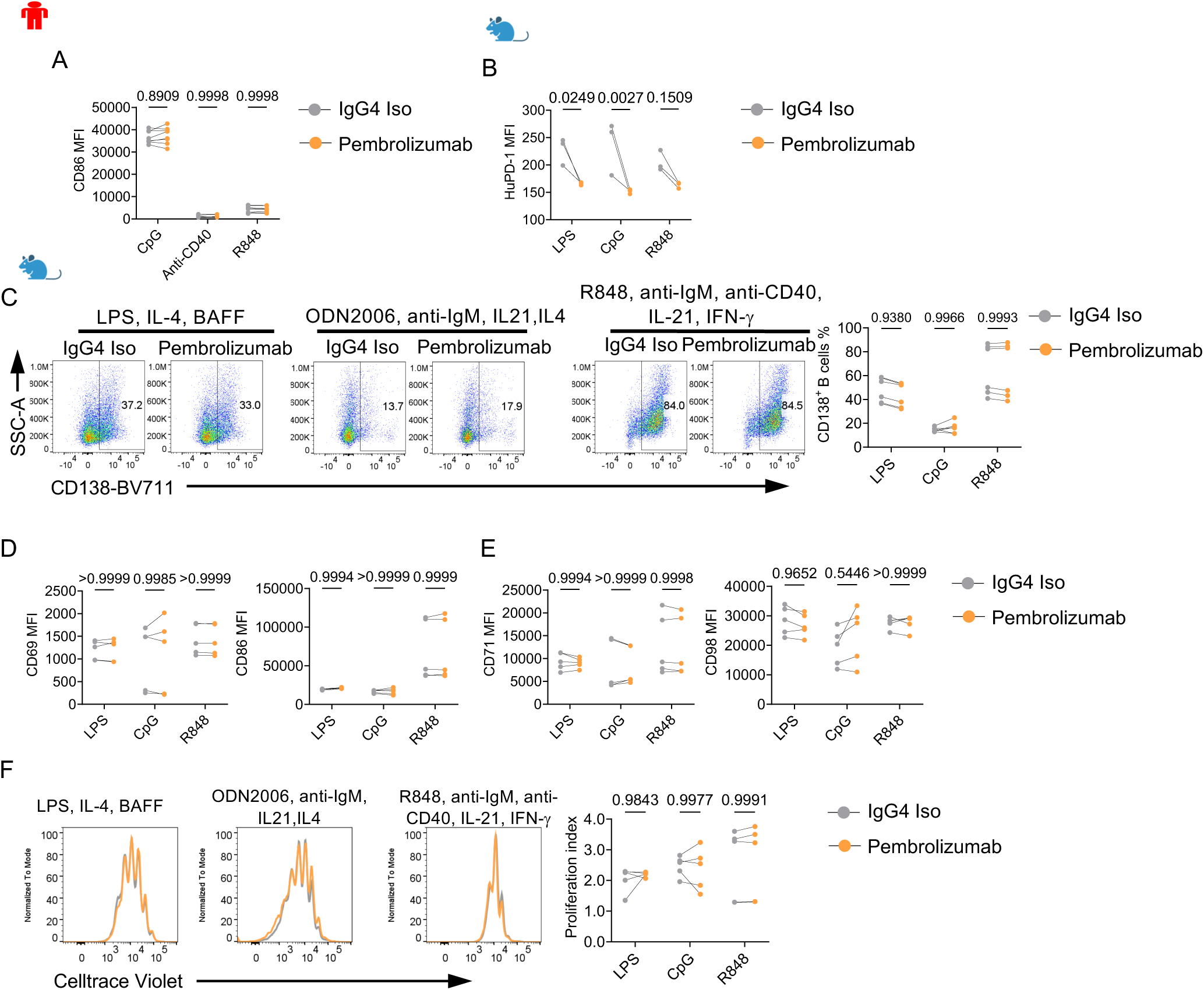
PD-1 inhibition does not have substantial impact on B cells *ex vivo* (A) Human naïve B cells were isolated from healthy donor PBMCs and stimulated with different conditions; Pembrolizumab (Keytruda) or isotype control IgG4 was added into the culture media on day 2, mean fluorescence intensity (MFI) of CD86 on B cells was measured, n = 8. (B-F) B cells were isolated from HuPD-1 mice, labeled with CTV, and cultured under 3 different conditions: LPS, rmIL-4, BAFF or Anti-IgM, CpG ODN, rmIL-21, rmIL-4, 100 rhIL-2 or R848, anti-CD40, anti-IgM, rmIL-21, rmIFN-γ for 3 days. Pembrolizumab or isotype control IgG4 was added on day 1, n = 5. (B) Summary of human PD-1 MFI on B cells from different groups. (C) Expression of CD138 on activated B cells. Right, percentage of CD138^+^ B cells. (D) Summaries of activation marker CD69 and CD86 MFIs on B cells from different groups. (E) Summaries of CD71 and CD98 MFIs on B cells from different groups. (F) Representative flow cytometry plot of CTV dilution on B cells. Right, a summary of the proliferation index of B cells from different groups. Data in graphs represent mean ± SEM, Significance was tested by Two-way ANOVA.

**Fig. S5.**
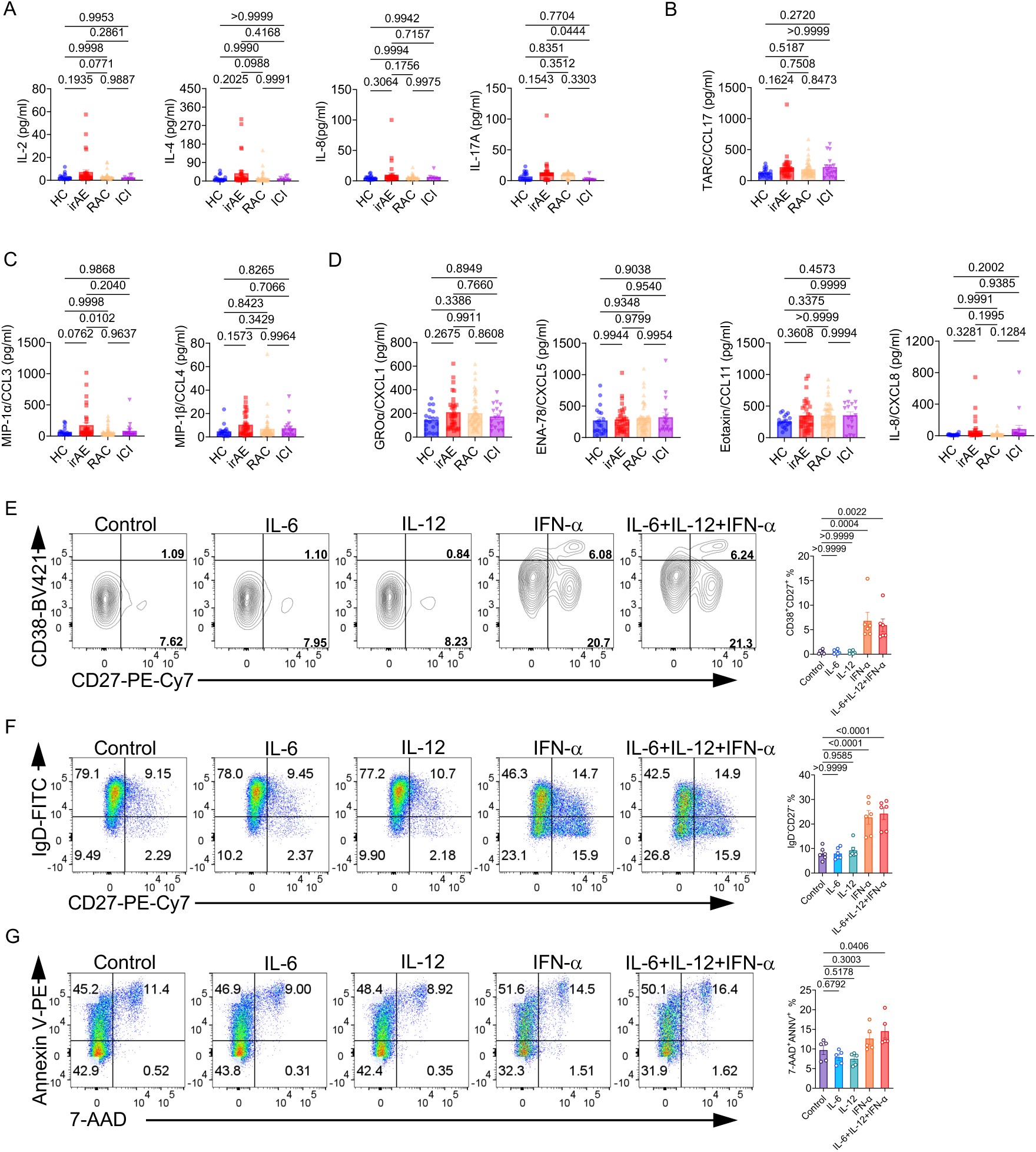
Inflammatory signatures enriched in irAE patients affect B cell activation and survival (A) Levels of inflammatory cytokines IL-2, IL-4, IL-8, and IL-17A in the plasma of patients, HC (n = 19), irAE (n = 34), RAC (n = 45), ICI (n = 9). (B-D) Levels of chemokine in the plasma, HC (n = 17), irAE (n = 33), RAC (n = 46), ICI (n = 17), CCL17 (B), MIP-1α and MIP-1β(C), CXCL1, CXCL5, CCL11, and CXCL8 (D). (E) Expression of CD27 and CD38 on B cells. Right, percentage of CD27^hi^CD38^hi^ ASCs, n = 6. (F) Expression of CD27 and IgD on B cells. Right, percentage of IgD^-^CD27^-^ B cells, n = 6. (G) Expression of Annexin V and 7-AAD on B cells. Right, percentage of 7-AAD^+^Annexin V^+^ B cells, n = 5. Data in graphs represent mean ± SEM, Significance was tested by One-way ANOVA.

**Fig. S6.**
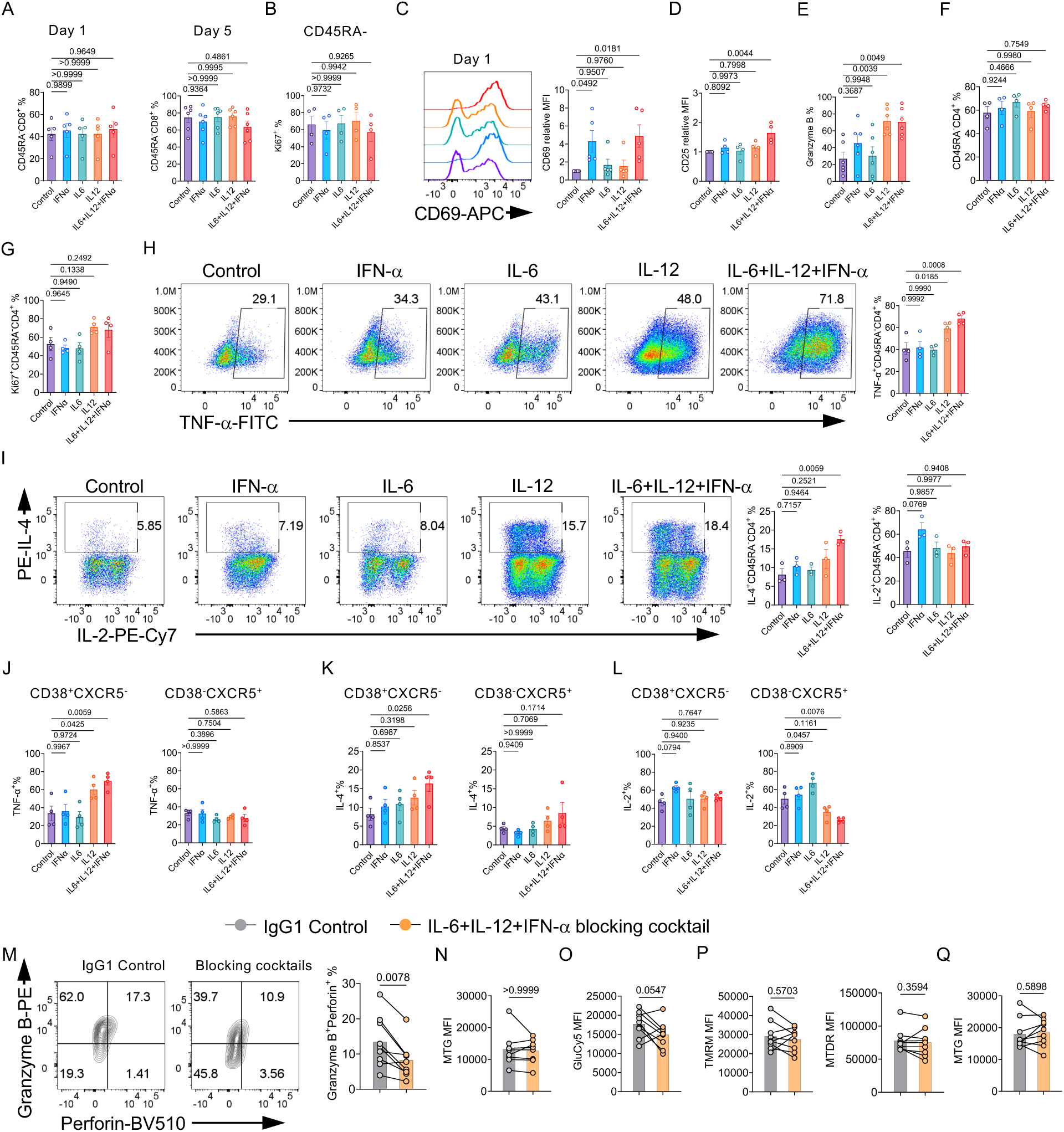
Inflammatory cytokines enriched in irAE patients promote both CD4^+^ and CD8^+^ T cell functions (A) Summaries of the percentage of CD45RA^-^CD8^+^ T cells at day 1 or day 5 of the culture, n = 6. (B) Summary of the percentage of Ki67^+^ in CD45RA^-^CD8^+^ T cells, n = 4. (C) Expression of CD69 on cultured CD8^+^ T cells at day 1. Right, CD69 mean fluorescence intensity (MFI) relative to the control in each experiment, n = 5. (D) Summary of the CD25 MFI relative to the control in each experiment, n = 5. (E) Summary of the percentage of Granzyme B^+^CD8^+^ T cells, n = 5. (F) Summary of the percentage of CD45RA^-^CD4^+^ T cells at day 5 of the culture, n = 4. (G) Summary of the percentage of Ki67^+^CD45RA^-^CD4^+^ T cells, n = 4. (H-L) CD4^+^ T cells were isolated from healthy donor PBMCs, stimulated with 10 μg/mL plate-coated anti-human CD3 and anti-human CD28 in the presence of vehicle control (control), 100 ng/mL IFN-α, 100 ng/mL IL-6, 100 ng/mL IL-12, or the combination of them for 5 days, followed by restimulating with PMA/Ionomycin/Monensin for 5 h. (H) Expression of TNF-α in CD45RA^-^CD4^+^ T cells. Right, percentage of TNF-α^+^ cells in CD45RA^-^CD4^+^ T cells, n =4. (I) Expression of IL-2 and IL-4 in CD45RA^-^CD4^+^ T cells. Right, percentage of IL-4^+^ and IL-2^+^ cells in CD45RA^-^CD4^+^ T cells, n = 3. (J-L) Summaries of the percentages of TNF-α^+^ (J), IL-4^+^ (K), or IL-2^+^ (L) cells in CD38^+^CXCR5^-^ or CD38^-^CXCR5^+^CD4^+^ T cells at day 5, n = 4. (M-Q) PBMCs from irAE patients were stimulated with 10 μg/mL plate-coated anti-human CD3 and anti-human CD28 in presence of IgG1 isotype control or a combination of 50 μg/mL anti-human IL-6R, 50 μg/mL anti-human IL-12p40 and 50 μg/mL anti-human IFNAR1 for 3 days, n = 9. (M) Expression of granzyme B and perforin in CD8^+^ T cells. Right, percentage of granzyme-B^+^perforin^+^CD8^+^ T cells. (N) MFI of MTG in CD8^+^ T cells. (O, P) MFI of GluCy5 (O), TMRM and MTDR (P), and MTG (Q) in CD4^+^ T cells. Data in graphs represent mean ± SEM, Significance was tested by One-way ANOVA (A-L), and paired Student’s T-test (N-Q).

**Supplementary Table 1.**
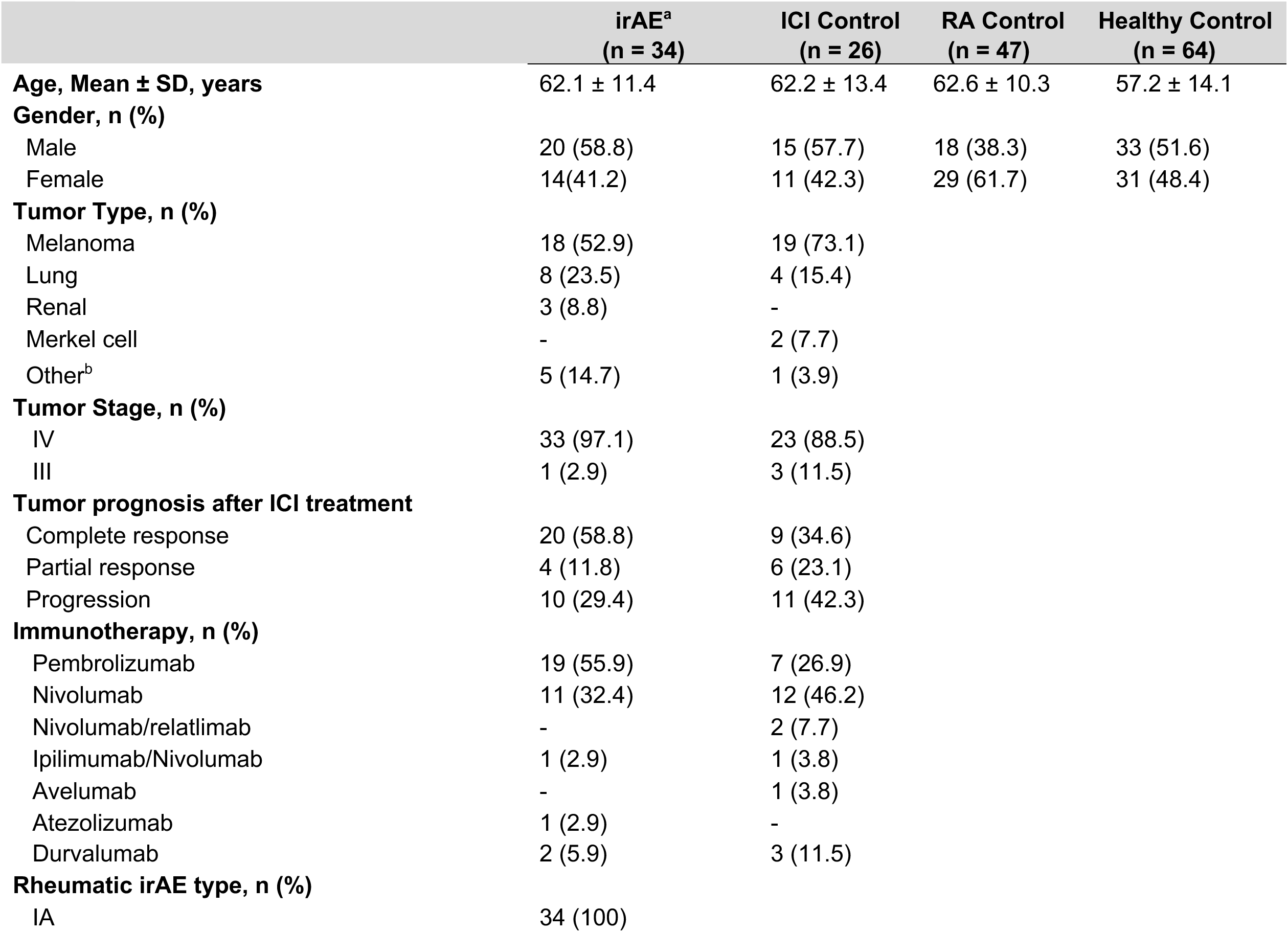
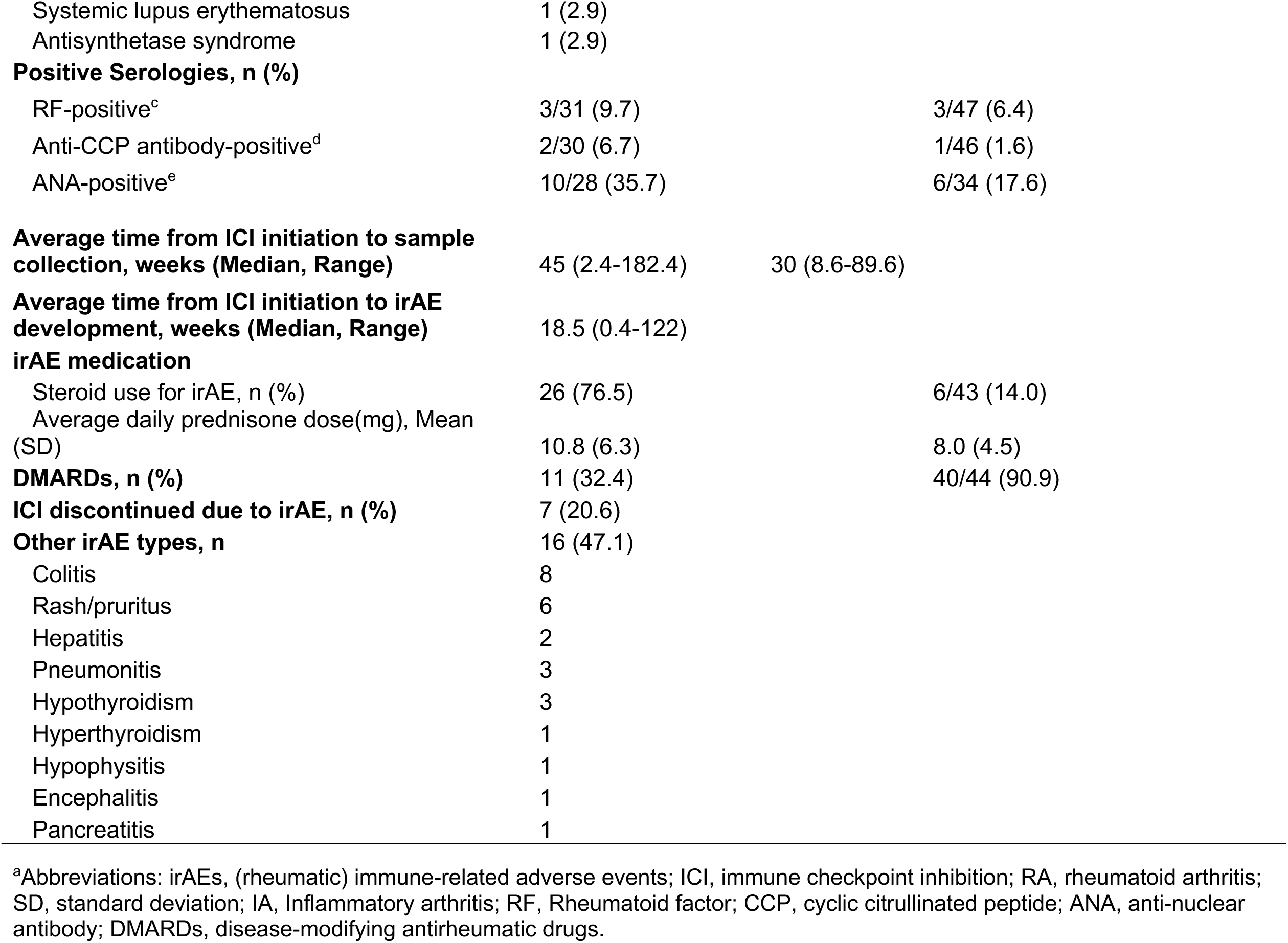
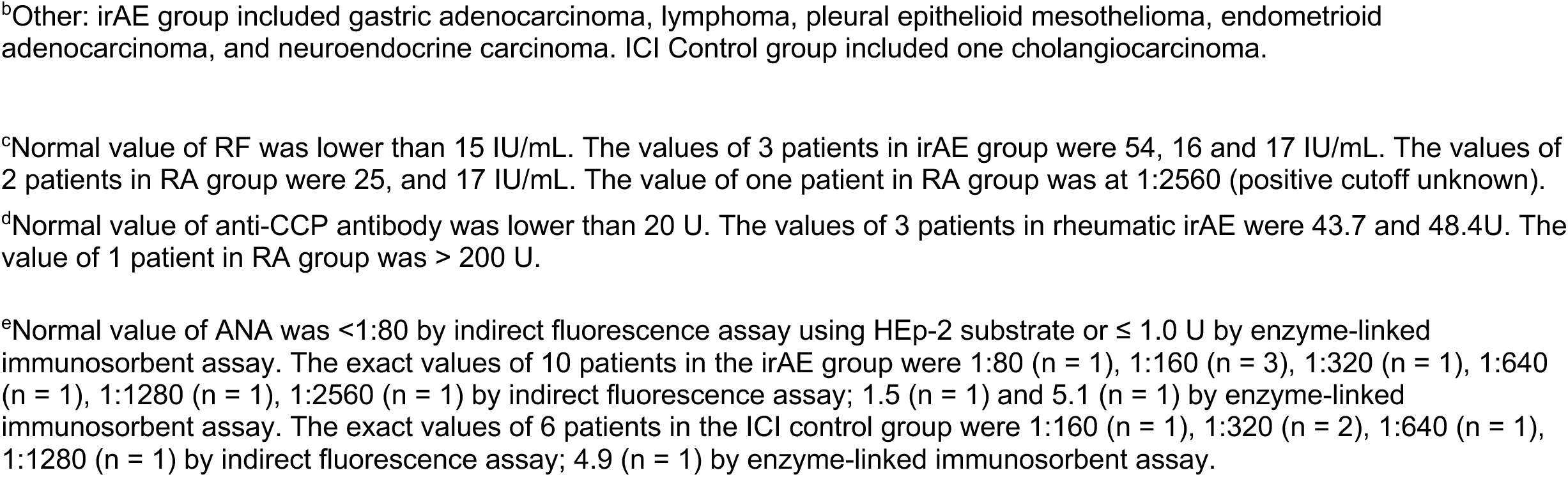
Patient demographics and disease characteristics.

